# Genetic regulation of *OAS1* nonsense-mediated decay underlies association with risk of severe COVID-19

**DOI:** 10.1101/2021.07.09.21260221

**Authors:** A Rouf Banday, Megan L Stanifer, Oscar Florez-Vargas, Olusegun O Onabajo, Muhammad A Zahoor, Brenen W Papenberg, Timothy J Ring, Chia-Han Lee, Evangelos Andreakos, Evgeny Arons, Greg Barsh, Leslie G Biesecker, David L Boyle, Andrea Burnett-Hartman, Mary Carrington, Euijin Chang, Pyoeng Gyun Choe, Rex L Chrisholm, Clifton Dalgard, Jeff Edberg, Nathan Erdmann, Heather S Feigelson, Gary S Firestein, Adam J Gehring, Michelle Ho, Steven Holland, Amy A Hutchinson, Hogune Im, Michael G Ison, Hong Bin Kim, Robert J Kreitman, Bruce R Korf, Lisa Mirabello, Jennifer A Pacheco, Michael J Peluso, Daniel J Rader, David T Redden, Marylyn D Ritchie, Brooke Rosenbloom, Hanaisa P Sant Anna, Sharon Savage, Eleni Siouti, Vasiliki Triantafyllia, Joselin M Vargas, Anurag Verma, Vibha Vij, Duane R Wesemann, Meredith Yeager, Xu Yu, Yu Zhang, Steeve Boulant, Stephen J Chanock, Jordan J Feld, Ludmila Prokunina-Olsson

## Abstract

Genomic regions have been associated with COVID-19 susceptibility and outcomes, including the chr12q24.13 locus encoding antiviral proteins OAS1-3. Here, we report genetic, functional, and clinical insights into genetic associations within this locus. In Europeans, the risk of hospitalized vs. non-hospitalized COVID-19 was associated with a single 19Kb-haplotype comprised of 76 *OAS1* variants included in a 95% credible set within a large genomic fragment introgressed from Neandertals. The risk haplotype was also associated with impaired spontaneous but not treatment-induced SARS-CoV-2 clearance in a clinical trial with pegIFN-λ1. We demonstrate that two exonic variants, rs10774671 and rs1131454, affect splicing and nonsense-mediated decay of *OAS1*. We suggest that genetically-regulated loss of *OAS1* expression contributes to impaired spontaneous clearance of SARS-CoV-2 and elevated risk of hospitalization for COVID-19. Our results provide the rationale for further clinical studies using interferons to compensate for impaired spontaneous SARS-CoV-2 clearance, particularly in carriers of the *OAS1* risk haplotypes.

## TEXT

The complex response to pathogens, such as SARS-CoV-2, the causative agent of COVID-19, is likely determined by the interplay between host and pathogen factors. Variability in clinical outcomes of COVID-19 has prompted the search for host genetic factors to elucidate underlying disease mechanisms and guide optimal treatment options. Recent genome-wide association studies (GWAS) have reported a series of genetic variants in distinct loci associated with COVID-19 susceptibility and increased risk of hospitalization or severe disease compared to the general population^1,2^.

The associated locus at 12q24.13^1,2^, marked by rs10774671, harbors three genes encoding antiviral 2’,5’-oligoadenylate synthetase (OAS) enzymes — OAS1, OAS2, and OAS3 — which are interferon-inducible effectors activating the latent form of ribonuclease L (RNaseL)^3,4^. Activation of the RNaseL pathway leads to degradation of viral RNA, inhibition of virus replication, and cell death^3^. A 185 Kb-haplotype spanning OAS1, OAS2, and OAS3 was reported as introgressed into genomes of modern humans from the Neandertal and Denisova lineages^5-7^. The risk of severe COVID-19 was associated with rs10774671-A allele capturing the non-Neandertal haplotype^1,2^. Alleles of rs10774671 affect splicing of 3’ terminal exons of *OAS1*. Specifically, the rs10774671-G allele creates a new splicing site for inclusion of exon 6, creating a transcript for a protein isoform of 46 KDa known as OAS1-p46. Other splicing sites are utilized in the absence of G allele (and presence of A allele), creating transcripts for other OAS1 isoforms, including OAS1-p42^8^. Notably, rs10774671-A allele was also associated with impaired spontaneous clearance of West Nile virus infection^9^ and decreased basal OAS enzyme activity in unstimulated peripheral blood mononuclear cells (PBMCs) from healthy individuals^8^. Additionally, rs10774671-A, as well as linked alleles of other variants in this region have been associated with chronic hepatitis C virus infection^10^, impaired SARS-CoV clearance^11^, and increased susceptibility to several autoimmune conditions - multiple sclerosis^12,13^, Sjögren’s syndrome^14^, and systemic lupus erythematosus^15,16^. A linked variant, rs4767027 (r^2^=0.97 with rs10774671 in Europeans), has been reported as a protein quantitative trait locus (pQTL) for OAS1 blood levels in the general population^17^. Because OAS1 levels were decreased in the presence of rs4767027-C allele (linked with the GWAS rs10774671-A risk allele), this suggested that OAS1 deficiency could underly the risk of severe COVID-19^17^. However, the factors regulating OAS1 expression were not explored.

Here, we sought to explore the underlying mechanism of the COVID-19 GWAS locus at 12q24.13. In case-case analyses in 1555 individuals of European ancestry, we refined the associations of the Neandertal and other haplotypes with COVID-19 severity. Further, we employed a series of bioinformatics and experimental approaches to demonstrate the combined effects of two associated variants on post-transcriptional regulation of *OAS1* expression through nonsense-mediated decay (NMD). By analysis in a clinical trial with recombinant interferon lambda (pegIFN-λ1)^18^, we demonstrated that the *OAS1* haplotype associated with an elevated risk of hospitalized COVID-19 was also associated with impaired spontaneous but not pegIFN-λ1-induced clearance of SARS-CoV-2. These results offer a molecular mechanism underlying the COVID-19 related genetic associations within the 12q24.13 locus and suggest the utility of further pursuit of treatment with pegIFN-λ1 or other interferons to compensate for deficiencies in spontaneous clearance in carriers of the *OAS1* risk haplotypes.

## RESULTS

### Case-case analyses within the 12q24.13 locus refine the associations with COVID-19 outcomes

We investigated the 116Kb region (hg38, chr12: 112,901,422-113,017,487) within the 12q24.13 locus, which includes *OAS1, OAS2*, and *OAS3* genes. In COVID-19 Host Genetics Initiative (HGI) GWAS meta-analysis^2^, rs10774671 (hg38, chr12:112919388-G-A) was reported as the lead variant in this region for associations with critical disease (A2 analysis, p=1.1E-12), hospitalized disease (B2, p=1.51E-09), and reported SARS-CoV-2 infection (C2, p=2.09E-11), all compared to the general population^2^. However, rs10774671 was not associated with hospitalized vs. non-hospitalized disease (B1, p=0.80) (release r5, COVID-19 HGI Browser https://app.covid19hg.org/).

We analyzed 366 genotyped or imputed variants from this region in an independent set of 1555 patients of European ancestry from COVNET. A 32Kb linkage disequilibrium (LD) block (r^2^>0.8 with rs10774671) spanning OAS1 and OAS3 included 125 variants associated with p=1.72E-04 to 4.22E-06 comparing all hospitalized vs. non-hospitalized patients (**Table S1, Figure 1a**). Applying the Sum of Single Effects (SuSiE) analysis^19^, we identified two credible sets, each predicted to include at least one effect variant with 95% certainty. The lead rs10774671 was included in a credible set of 76 variants, all in r2>0.9 with the lead rs10774671 and spanning OAS1; the second credible set included 12 OAS3 variants in low LD with the first set, possibly indicating an independent but weaker signal (**Table S1, Figure 1b**).

**Figure 1.**
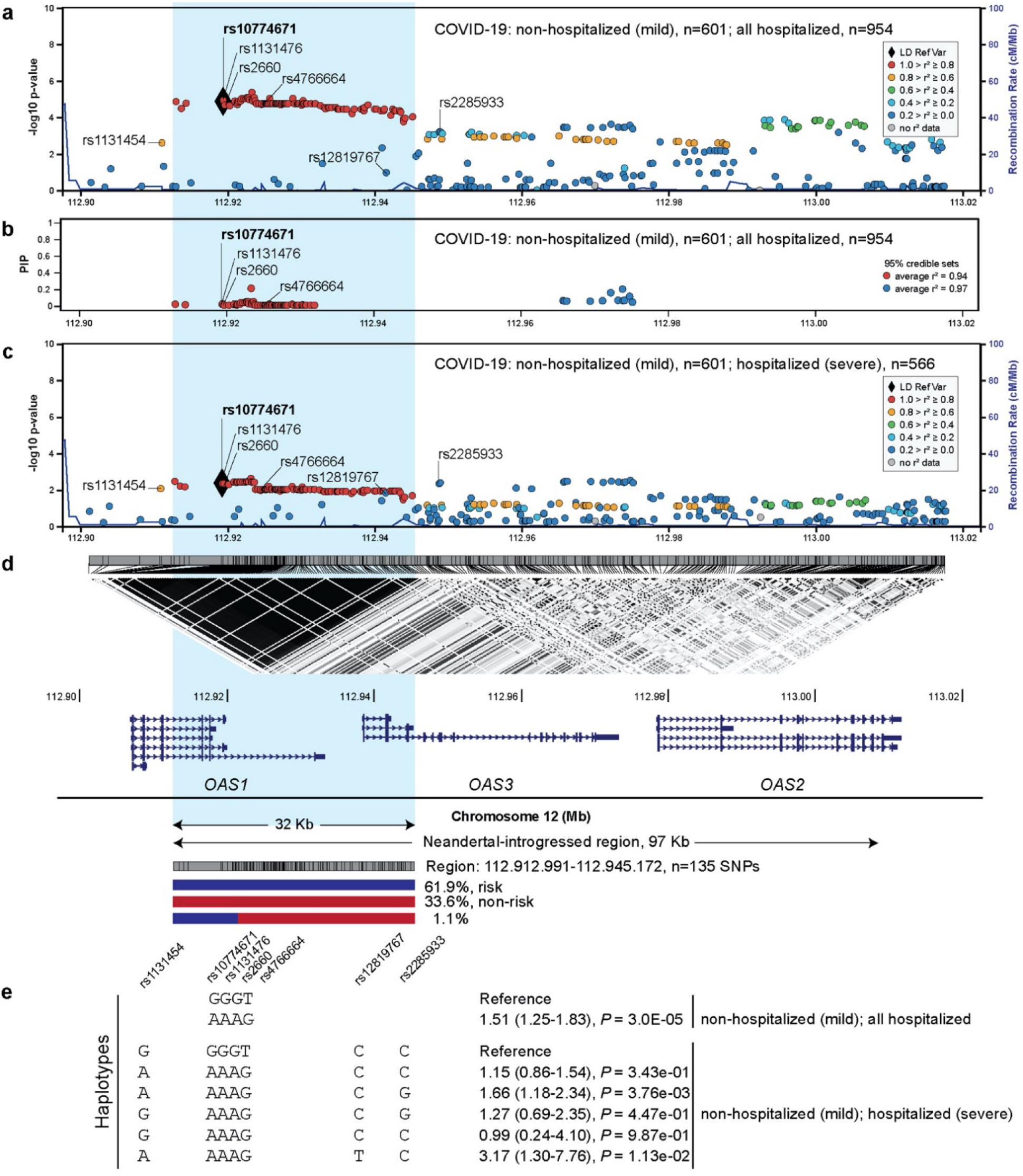
Association analyses within the chr12q24.13 region in relation to COVID-19 outcomes in patients of European ancestry. **a)** Association analyses in 601 patients with non-hospitalized (mild) compared to 954 patients with hospitalized COVID-19. **b)** Variants included in SuSie 95% credible sets and their posterior inclusion probabilities (PIP); two 95% credible sets were identified with a threshold of an average correlation between all variables >0.9. **c)** Association analyses in 601 patients with non-hospitalized (mild) compared to 566 patients with hospitalized severe COVID-19. d) LD plot (r^2^) in the total set of 1555 patients with COVID-19 and location of *OAS1, OAS3*, and *OAS2* reference transcripts. Blue shading indicates the 32Kb LD block (hg38: chr12:112912991-112945172) that includes 135 variants in r^2^>0.8 with the lead rs10774671; 125 of these variants form only two common haplotypes – risk and non-risk. The Neandertal-introgressed genomic fragment of 97 Kb is marked based on previously reported variants^7^. **e)** Analysis of haplotypes including four core variants tagging the Neandertal haplotype and extended haplotypes additionally including three variants associated with severe COVID-19. Full association results for individual variants and haplotypes are provided in **Table S1**.

A similar analysis limited to 1167 patients with hospitalized severe vs. non-hospitalized disease identified an additional set of 24 variants with suggestive associations (p=0.045-0.0049, **Table S1, Figure 1c**). This set included three missense variants in moderate or low LD with rs10774671: an OAS1-exon 3 rs1131454 (Gly162Ser, OR=1.36 (1.07-1.74, p=0.014, r^2^=0.73 with rs10774671), a common OAS3-exon 6 rs2285933 (Ser381Arg, OR=1.47 (1.11-1.94), p=0.0066, r^2^=0.18) and an uncommon European-specific OAS3-exon 2 rs12819767 (Arg65Trp, OR=2.73 (1.13-6.52), p=0.024, r^2^=0.009). The SuSiE analysis was not informative for this outcome due to less significant associations precluding generation of 95% credible sets (data not shown).

The 125 variants within the OAS1/OAS3 LD block formed only two common haplotypes – the risk (61.9%) and non-risk (33.6%). The non-risk haplotype included alleles defining the Neandertal haplotype^20^, which spans >97Kb of this region (**Figure 1d**). Our OAS1 credible set of 76 variants further limited the potentially causal region of this haplotype to 19Kb within OAS1 (**Table S1A**). From this credible set, we selected four directly genotyped variants spanning a 4.2Kb region (rs10774671, rs1131476, rs2660, and rs4766664) and capturing the Neandertal haplotype in Europeans. In a comparison to the Neandertal haplotype (GGGT), there was only one common risk haplotype formed by these variants (AAAG, OR=1.51 (1.25-1.83), p=3E-05, n=1555) (**Table S2, Figure 1e**).

Analyses of the extended haplotypes (adding three additional variants associated with severe disease to the core four-variant haplotype) demonstrated that rs12819767-T and rs2285933-G alleles increased and rs1131454-G allele decreased the associations with severe disease of the core risk haplotype (AAAG, **Table S3, Figure 1e**). In smaller sets of patients of non-European ancestries (483 African, 166 admixed-Hispanic, and 103 East-Asian), none of these variants or their haplotypes showed significant associations (**Table S4-S8**). Thus, in Europeans, the risk of hospitalized disease was largely associated with one non-Neandertal haplotype comprising a 95% credible set of 76 OAS1 variants, while investigation in other populations awaits larger studies. Additional OAS1 and OAS3 missense variants and their corresponding haplotypes might be further contributing to the association with severe COVID-19.

### OAS1 allelic protein isoforms have comparable anti-SARS-CoV-2 activity

Our analyses suggested that in Europeans at least one effect variant for the risk of hospitalized disease is included in the 95% credible set of 76 *OAS1* variants. This set included four missense variants (rs1131476, rs1051042, rs2660, and rs11352835) and one splicing variant (rs10774671). Additionally, an *OAS1* missense variant rs1131454 was included in a haplotype associated with severe disease. The alleles of rs10774671 create distinct protein isoforms OAS1-p42 (A-allele) and OAS1-p46 (G-allele)^8^, but the functional effects of other associated *OAS1* variants, especially in the context of OAS1 protein isoforms, have not been explored. To test the anti-SARS-CoV-2 activity of OAS1 proteins generated by the risk and non-risk haplotypes, we generated four expression constructs corresponding to OAS1-p42 and p46 proteins (**Figure 2a**). We transiently overexpressed all OAS1 plasmids in a lung epithelial cell line A549-ACE2 stably expressing ACE2, the SARS-CoV-2 receptor. To control for endogenous interferon response to the plasmids, we transfected cells either with a control GFP plasmid or individual *OAS1* plasmids and, after 48 hours, infected cells with SARS-CoV-2 for 24 hours (**Figure 2b**). The A549-ACE2 cells were highly infectable by SARS-CoV-2, but viral loads were similarly decreased by overexpression of all *OAS1* plasmids (**Figure 2c, Table S9**). Thus, in A549-ACE2 cells, proteins produced by similar amounts of OAS1 plasmids provided comparable anti-SARS-CoV-2 activity, without a detectable functional impact of the splicing (rs10774671) or missense (rs1131454, rs1131476, and rs1051042) variants (**Figure 2c**).

**Figure 2.**
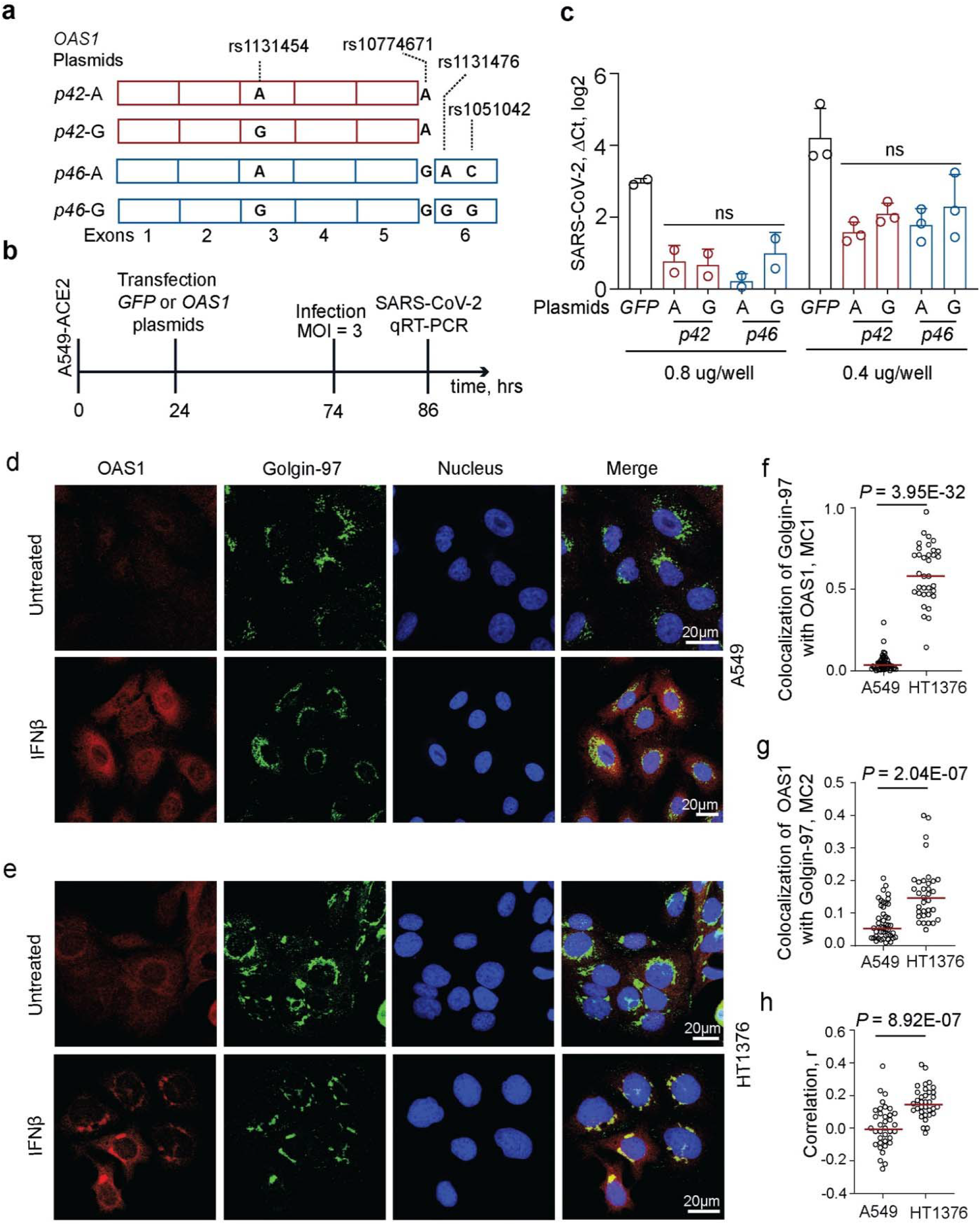
Anti-SARS-CoV-2 activity and subcellular localization of OAS1-p42 and p46 isoforms. **a)** Description of *OAS1-p42* and *OAS1-p46* plasmids. **b)** Experiment outline: plasmids were transiently transfected in A549-ACE2 cells, followed by infection with SARS-CoV-2 and qRT-PCR for viral detection. c) SARS-CoV-2 load in A549-ACE2 cells transfected in duplicates or triplicates with 0.4 ug/well or 0.8 ug/well with *OAS1* or *GFP* plasmids in six-well plates. Expression of SARS-CoV-2 was detected by qRT-PCR and normalized to the expression of an endogenous control (*HPRT1*). P-values between *OAS1* plasmids are for ANOVA tests. **d, e)** Representative confocal images for endogenous OAS1 expression in untreated and IFNβ-treated A549 cells (rs10774671-AA, OAS1-p42, cytosolic expression) and HT1376 (rs10774671-GG, OAS1-p46, enrichment in trans-Golgi compartment); OAS1 (red), Golgin-97 (green) and nuclei (DAPI, blue). Scale bars, 20⍰µm. **f)** Mander’s coefficient 1 (MC1) for colocalization of Golgin-97 with OAS1 in confocal images. **g)** Mander’s coefficient 2 (MC2) for colocalization of OAS1 with Golgin-97 in confocal images. **h)** Overall correlation (Pearson’s, r) between colocalization of Golgin-97 and OAS1 expression in confocal images. P-values are for non-parametric, two-sided Mann– Whitney U tests.

OAS1-p42 and OAS1-p46 proteins have been shown to localize to the cytosol and the trans-Golgi compartment, respectively^21^. By confocal imaging of endogenous OAS1 proteins in IFNβ-treated cells, we confirmed that OAS1-p46 produced in HT1376 cells (rs10774671-GG) was enriched in the trans-Golgi compartment. In contrast, OAS1-p42 produced in A549 cells (rs10774671-AA) appeared exclusively cytosolic (**Figure 2d-h**). Despite differences in intracellular localization of OAS1 protein isoforms, determined by the splicing variant rs10774671, our results do not support their differential anti-SARS-CoV-2 activity.

### *OAS1* expression is regulated by a splicing enhancer created by missense variant rs1131454

The risk rs10774671-A allele was associated with lower OAS1 blood levels in the general population^17^. To explore whether this difference can be detected on mRNA level as well, we quantified the expression of *OAS1* isoforms in various RNA-seq datasets. Expression of *OAS1*-*p46* (created by rs10774671-G allele) was higher than of other *OAS1* isoforms in all datasets tested - adjacent normal tissues from TCGA (**Figure S1**), nasal epithelial cells infected with Rhinovirus (**Figure S2a**), pulmonary alveolar type 1 cell-based organoids infected with SARS-CoV-2 (**Figure S2b**), and in PBMCs from COVID-19 patients (**Figure S2c**).

Higher expression of the *OAS1-p46* isoform could be due to its enhanced transcription regulated by rs10774671 or its linked variants. However, based on analyses of chromatin profiles in three cell lines by ATAC-seq and H3K27ac ChIP-seq or chromatin interactions by Hi-C, we did not observe evidence for regulatory elements within the associated region (**Figure S3**). Similarly, we did not detect chromatin interactions involving this region by Hi-C in a monocytic cell line THP-1 at baseline or after treatment with IFNβ (**Figure S4**). Therefore, we excluded transcriptional regulation of *OAS1* expression by rs10774671 or its linked variants and considered post-transcriptional mechanisms.

As an indication of a potential imbalance in transcript production or stability, we evaluated allele-specific expression of transcribed *OAS1* variants. Specifically, we analyzed heterozygous samples from nasal epithelial cells – mock or in vitro infected with Rhinoviruses, and PBMCs from COVID-19 and non-COVID-19 patients. Using allele-specific RNA-seq reads, we analyzed two transcribed variants linked with rs10774671 - rs2660 in exon 6 (r^2^ =0.96 in Europeans, included in *OAS1*-*p46* and *p48* transcripts) and rs1131454 in exon 3 (r^2^=0.72 in Europeans, included in all *OAS1* transcripts). For both variants, the counts of RNA-seq reads with G alleles were significantly higher than reads with A alleles (**Figure 3a**). In most cases, rs1131454-G is linked with the rs10774671-G allele and included within the Neandertal haplotype. However, rs1131454-G also exists in the less common haplotype with the risk rs10774671-A allele. We analyzed samples heterozygous for rs1131454 (AG) but homozygous for the risk alleles of rs10774671 (AA) and rs2660 (AA). Analysis of haplotypes of these three variants in different datasets showed higher expression in samples with the GAA compared to AAA haplotypes, indicative of allele-specific expression for rs1131454. The expression of both *OAS1-p46* and *OAS1-p42* transcripts was increased in the presence of exon 3 rs1131454-G allele (**Figure 3b-f**).

**Figure 3.**
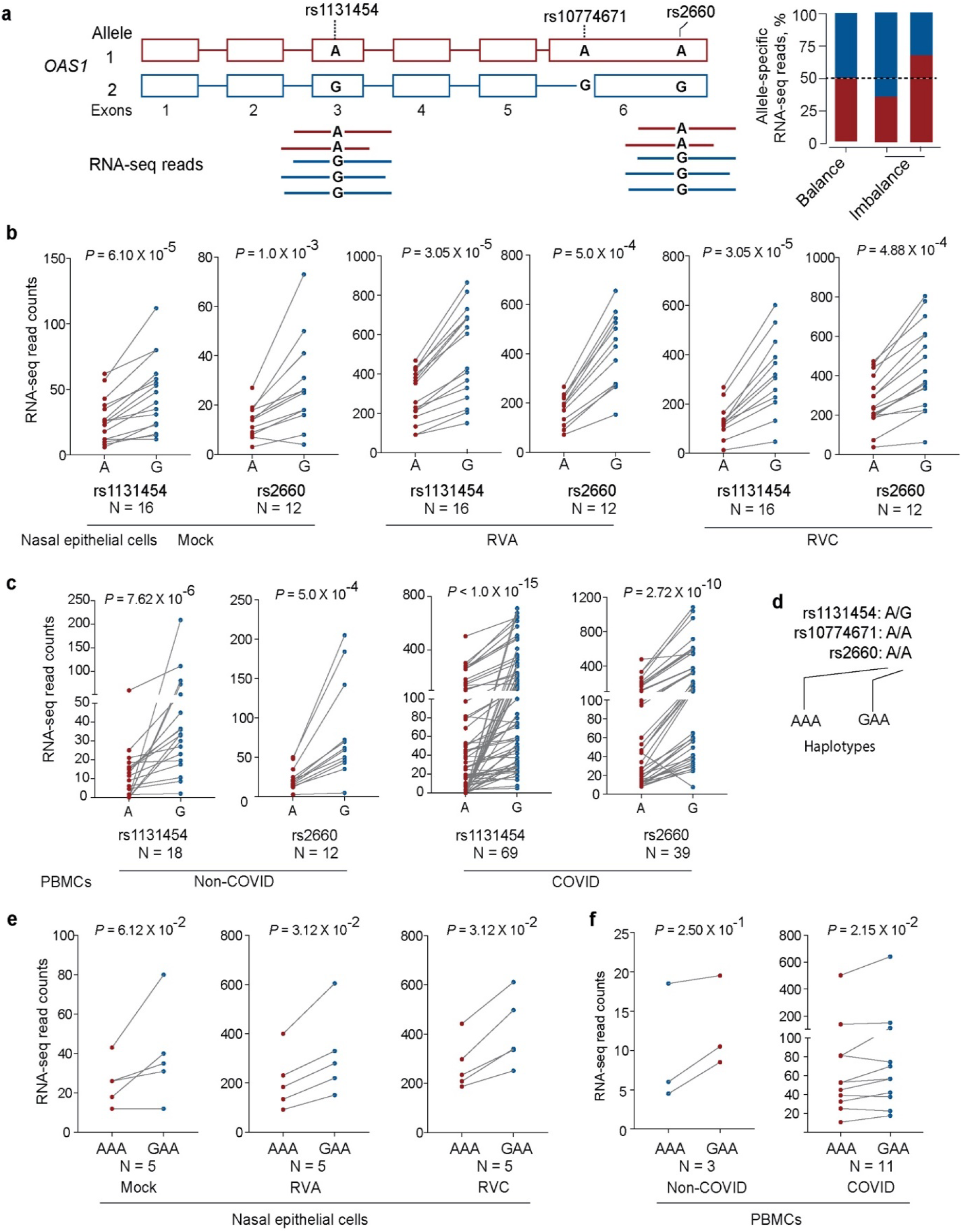
Allelic expression imbalance of *OAS1* transcripts. **a)** Outline of allelic imbalance analysis for transcribed *OAS1* variants based on RNA-seq reads. Counts of allele-specific RNA-seq reads in heterozygous samples for transcribed *OAS1* variants rs1131454 and rs2660 in **b)** nasal epithelial cells – uninfected (Mock) and infected with Rhinovirus (RV) strain A or C or **c)** in PBMCs from patients with COVID-19 and healthy controls (labeled as non-COVID). **d)** Haplotypes of the three *OAS1* variants used for analysis. **e)** Haplotype-specific imbalance in *OAS1* expression contributed by rs1131454. *P*-values are for non-parametric, Wilcoxon matched pairs signed-rank tests.

By visual examination of RNA-seq plots, we noted different lengths of *OAS1* exon 3 (labeled as Short and Long, **Figure 4a**), created due to alternative splicing through cryptic acceptor (∼15-25 % reads) and donor (∼5% reads) splice sites. Splice-QTL analyses showed increased canonical splicing and reduced alternative splicing of exon 3 in the presence of the rs1131454-G allele (**Figure 4b-d**). By *in-silico* analysis, we predicted that the rs1131454-G allele creates a putative exonic splicing enhancer/silencer (ESE/ESS) (**Figure 5a**). To test this prediction, we generated allele-specific mini-genes with the A or G alleles of rs1131454 (**Figure 5b**). By RT-PCR in two cell lines (A549 and T24), we detected increased splicing of Long exon 3 in the mini-gene with the non-risk rs1131454-G allele (**Figure 5c, d**). Based on these results, we conclude that the functional effect of the missense variant rs1131454 could be related to creating an allele-specific ESE/ESS regulating splicing of OAS1 Short and Long exon 3 isoforms.

**Figure 4.**
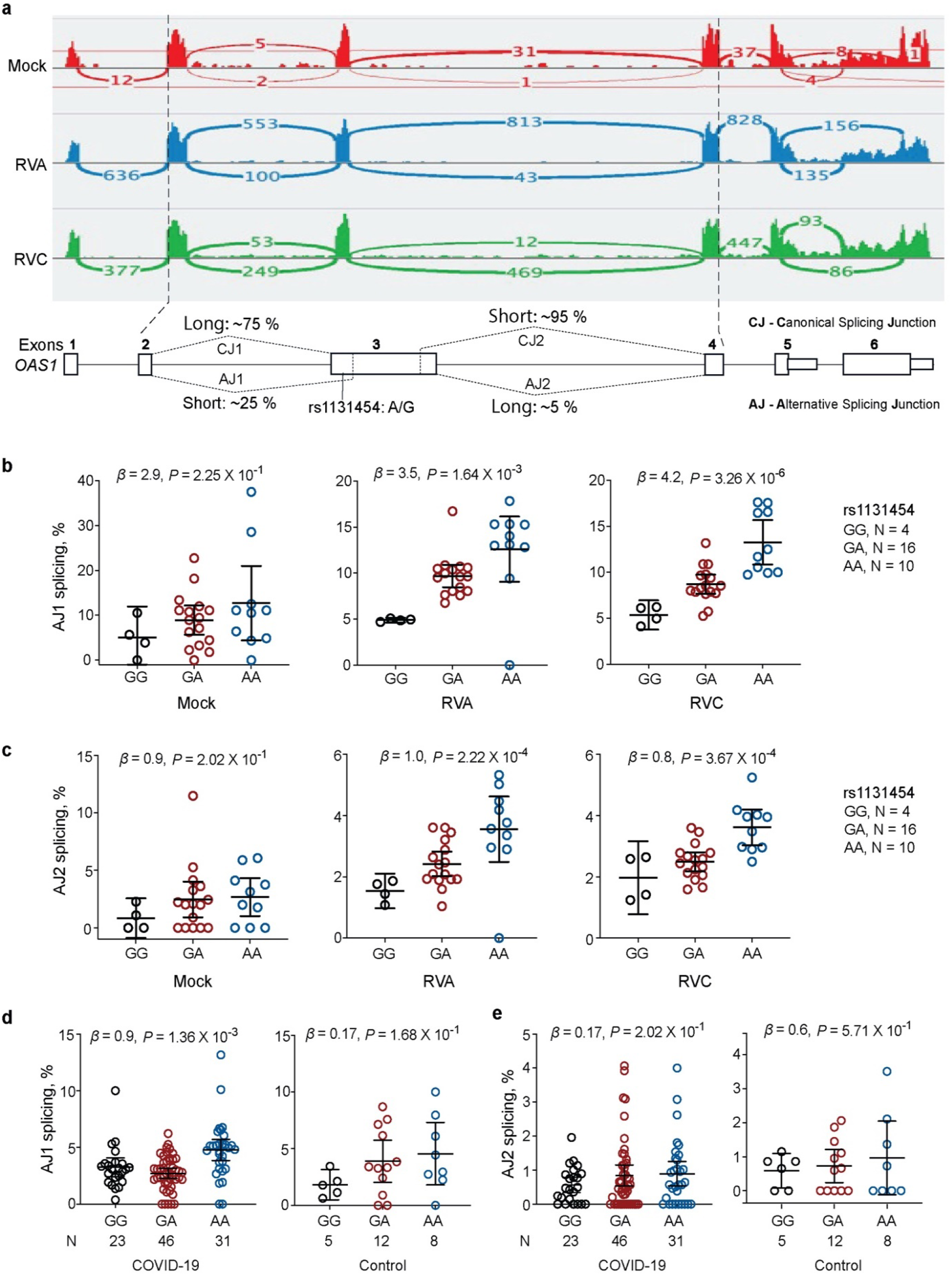
Splicing of *OAS1* exon 3 is associated with rs1131454. **a)** RNA-seq plots showing splicing patterns of *OAS1* exons in representative samples from nasal epithelial cells - uninfected (Mock) and Rhinovirus (RV)-infected with strains A and C. *OAS1* exon 3 shows alternative splicing at both 5’ acceptor and 3’ donor splice sites, resulting in four splicing junctions – two canonical junctions (CJ) and two alternative junctions (AJ) producing Long and Short versions of exon 3. Exon junctions AJ1 (major) and AJ2 (minor) account for approximately 25% and 5% of total RNA-seq reads, respectively. Splice quantitative trait locus (sQTL) analysis of AJ1 and AJ2 with rs1131454 in **b, c)** nasal epithelial cells – uninfected (Mock) or infected with RVA and RVC and **d, e)** in PBMCs from COVID-19 patients and healthy controls (labeled as non-COVID). *P*-values are for linear regression adjusting for sex and age. All graphs show individual data points with error bars representing 95% confidence intervals for the medians.

**Figure 5.**
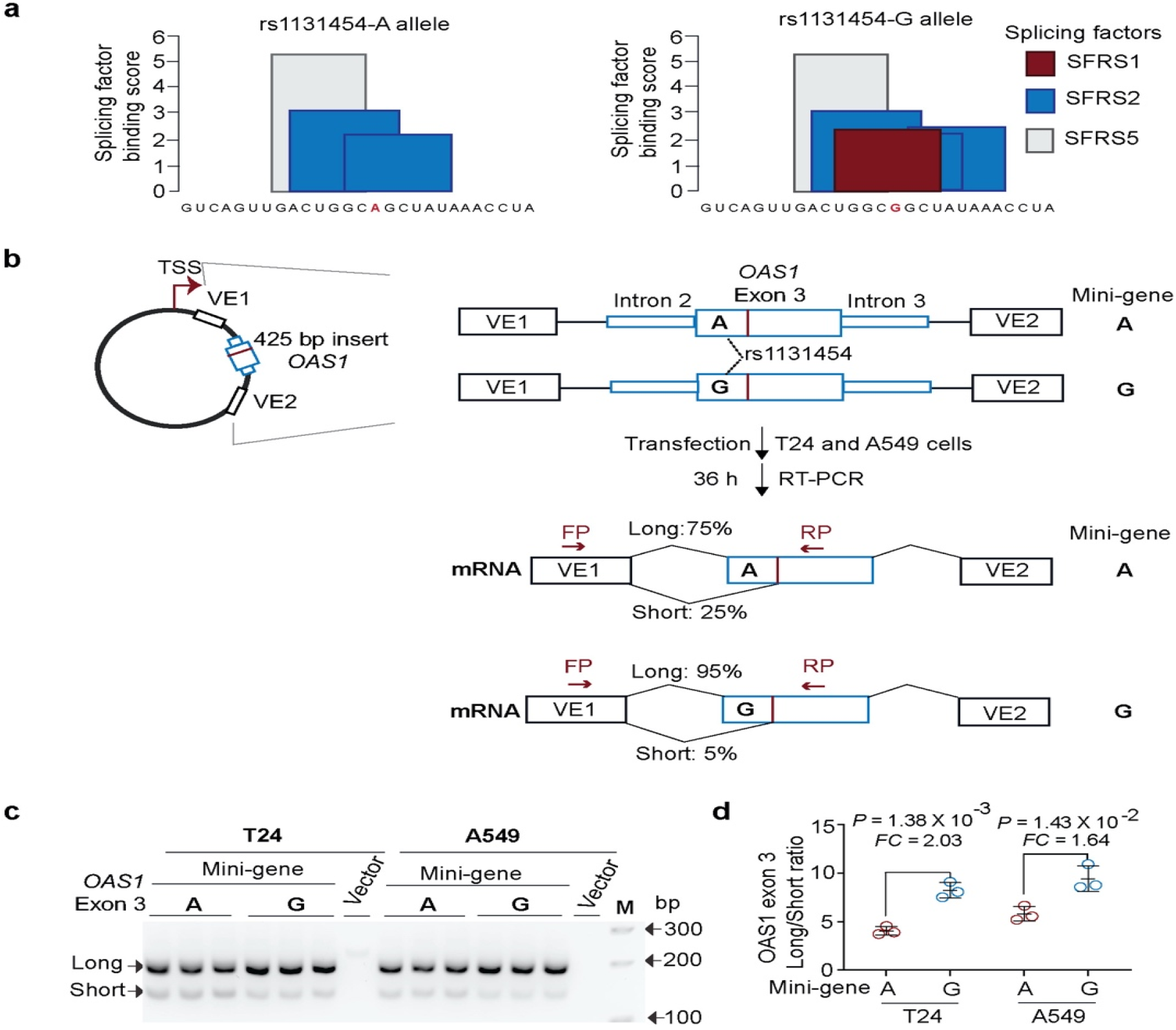
Exontrap assays demonstrate the functional effect of rs1131454 on *OAS1* exon 3 splicing. **a)** *In silico* prediction of allele-specific splicing factor binding sites within *OAS1* exon 3. Only the rs1131454-G allele creates a binding site for SFRS1 splicing factor; binding sites for SFRS2 are created by both alleles, with three or two sites in the presence of G or A allele, respectively. **b)** Experiment outline: description of allele-specific mini-genes with *OAS1* exon 3 inserts, transfection in T24 and A549 cells, and splicing ratios of amplicons detected by RT-PCR with FP and RP primers. **c)** A representative agarose gel showing splicing events of mini-genes detected by RT-PCR in T24 and A549 cells; Vector corresponds to negative control; M corresponds to 100-bp size marker. Upper and lower bands correspond to Long and Short exon 3 splicing events with vector exon 1 (VE1). No alternative splicing event was identified between exon 3 insert and VE2 (data not shown). Each mini-gene was analyzed in three biological replicates and the results of one of two independent experiments are shown. **d)** The ratios of Long/Short *OAS1* exon 3 expression quantified by densitometry of agarose gel bands. Splicing of Long exon 3 is significantly higher from the mini-gene with rs1131454-G allele compared to mini-gene with rs1131454-A allele. Fold changes (FC) were calculated from the splicing ratios. The dot plots are presented with means and SD; *P*-values are for unpaired, two-sided Student’s *t*-tests.

### *OAS1* transcripts are eliminated by allele-specific nonsense-mediated decay

To explore the impact of exon 3 splicing on *OAS1* transcripts, we performed long-read Oxford Nanopore RNA-sequencing in A549 and HT1376 cells at baseline and after treatment with IFNβ (**Figure S5**). Reads with alternatively spliced exon 3 were included in all *OAS1* transcripts (**Figure S6**). The combination of *p42* and *p46* isoforms and Short and Long exon 3 created several transcripts; all of these transcripts, except for *p46*-Long exon 3 isoform, had premature termination codons (PTCs, **Figure S6**). Prematurely terminated transcripts might be targeted by nonsense-mediated decay (NMD). To test this, we analyzed RNA-seq data for HeLa cells (rs10774671-AA, *OAS1-p42*) after siRNA-mediated knockdown (KD) of NMD pathway genes – *SMG6* and *SMG7*. The siRNA-KD of *SMG6* and *SMG7* resulted in upregulation of both alternative (Short) and canonical (Long) isoforms of exon 3, confirming that all *OAS1-p42* transcripts are targeted by NMD (**Figure 6a**,**b**).

**Figure 6.**
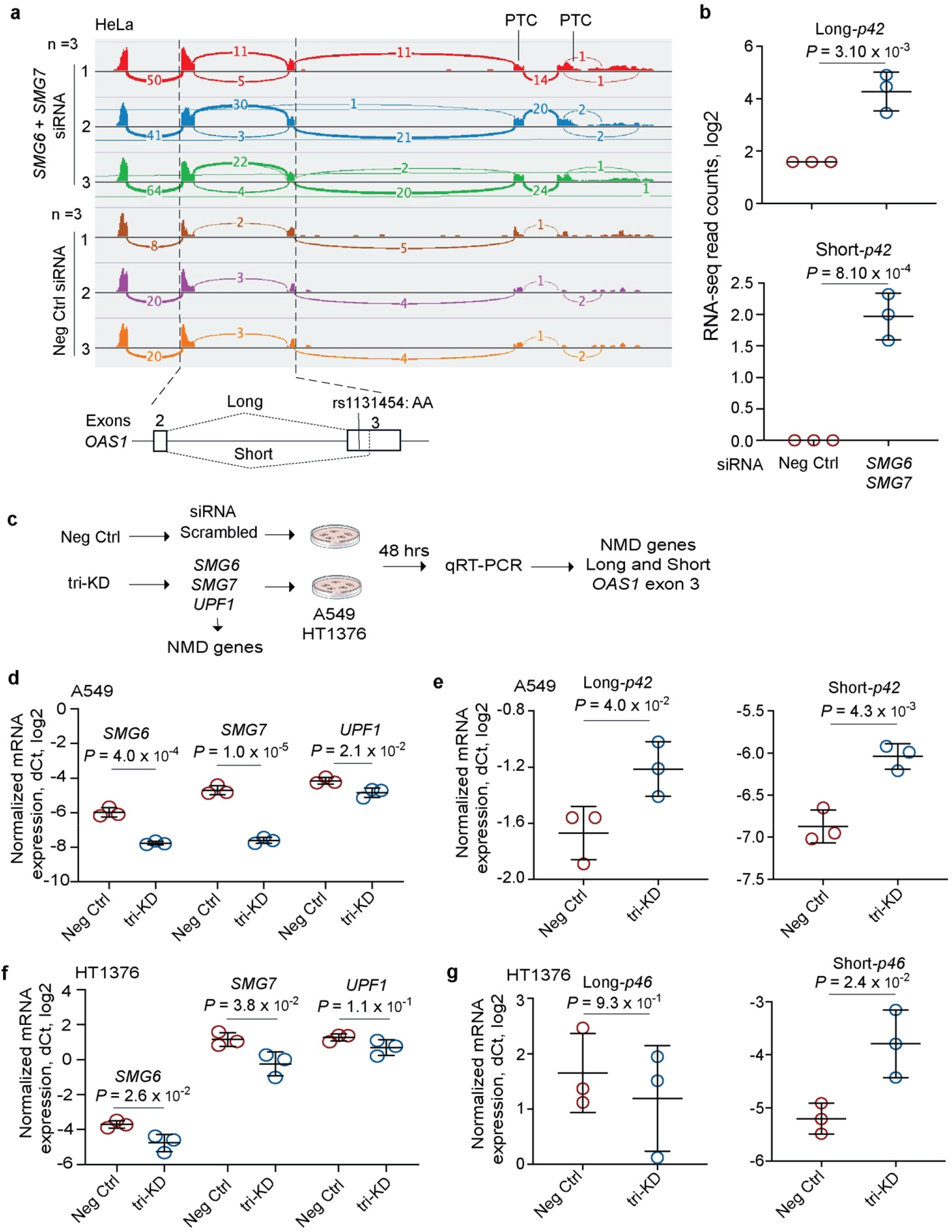
Nonsense-mediated decay targets *OAS1* isoforms with Short exon 3 upregulated by risk rs1131454-A allele. **a)** RNA-seq plots showing *OAS1* expression and splicing patterns in HeLa cells (rs10774671-AA, *OAS1-p42*) targeted by siRNA-knockdown (KD) of NMD-pathway genes SMG6 and SMG7 or by scrambled siRNA (Neg Ctrl). **b)** Expression of both Long and Short *OAS1*-*p42* isoforms is increased in HeLa cells with siRNA-KD of NMD genes. **c)** Schematics for characterizing effects of KD of NMD genes on the expression of Long and Short exon 3 in context of *OAS1-p42* (in A549) and *OAS1-p46* (in HT1376). **d, e)** Downregulation of NMD-pathway genes (*SMG6, SMG7* and *UPF1*) targeted by siRNA-KD in A549 cells. Expression of both Long and Short exon 3 *OAS1-p42* isoforms is increased in cells with siRNA-KD of NMD genes. **f, g)** Downregulation of NMD-pathway genes (*SMG6, SMG7* and *UPF1*) targeted by siRNA-KD in HT1376 cells. Only expression of Short-exon 3 *OAS1-p46* isoform is increased by siRNA-KD of NMD genes. Expression was analyzed by qRT-PCR and normalized to an endogenous control (*HPRT1*). The dot plots present means with SD; *P*-values are for unpaired, two-sided Student’s *t*-tests.

To test whether increased expression of *OAS1-p46*-Long could be due to its protection from NMD, we performed siRNA-KD of *SMG6, SMG7*, and UPFI (another component of the NMD pathway) in two cell lines - A549 (*OAS1-p42*) and HT1376 (*OAS1-p46*) and analyzed mRNA expression of Long and Short exon 3 as a readout (**Figure 6c**). In A549 cells, the triple-KD resulted in a significant increase in expression of both the Short and Long exon 3 versions of the *OAS1-p42* isoform (**Figure 6d**,**e**). However, in HT1376 cells, we observed a modest but significant increase of expression only of *OAS1-p46*-Short but not *OAS1-p46*-Long isoform (**Figure 6f,g**). We conclude that, unlike all other isoforms, the non-risk *OAS1-p46*-Long isoform is NMD-resistant. Because *OAS1* isoforms are created by haplotypes with rs1131454 and rs10774671 alleles, their combinations can contribute to the variation in NMD of *OAS1* transcripts.

### *OAS1* haplotypes are associated with impaired spontaneous clearance of SARS-CoV-2 that can be compensated by interferon treatment

Increased NMD of *OAS1-p42* transcripts, especially those with rs1131454-A allele, will result in significantly reduced OAS1 baseline expression levels. Early interferon signaling is important for mounting an efficient antiviral response^22^. For some infections, such as with SARS-CoV-2, that either do not induce sufficient interferon response or use various ways to counteract it, this deficit in the magnitude or delayed timing of the response could be crucial^23-26^. Interferon treatment has been proposed for mitigating SARS-CoV-2 infection, especially at an early infection stage^27,28^. To explore whether interferon treatment could prevent or mitigate infection *in vitro*, we treated infection-permissive intestinal cells Caco2^29,30^ (heterozygous both for rs10774671 and rs1131454) with IFNβ or IFN-λ cocktail four hours before or after SARS-CoV-2 infection. All treatments significantly decreased viral loads (**Figure 7a,b, Table S10**). In the same cells, *OAS1* expression was only minimally induced by SARS-CoV-2 alone but was strongly induced by pre- and post-treatment with both interferons (**Figure 7c, Table S11**). Interferon-induced expression of both OAS1-p42 and p46 isoforms was also detectable by Western blotting (**Figure S7**). These results suggested that interferons can induce robust immune response even in SARS-CoV-2 infected cells, inducing expression of both OAS1-p42 and OAS1-p46 isoforms and compensating for any loss of their expression due to NMD.

**Figure 7.**
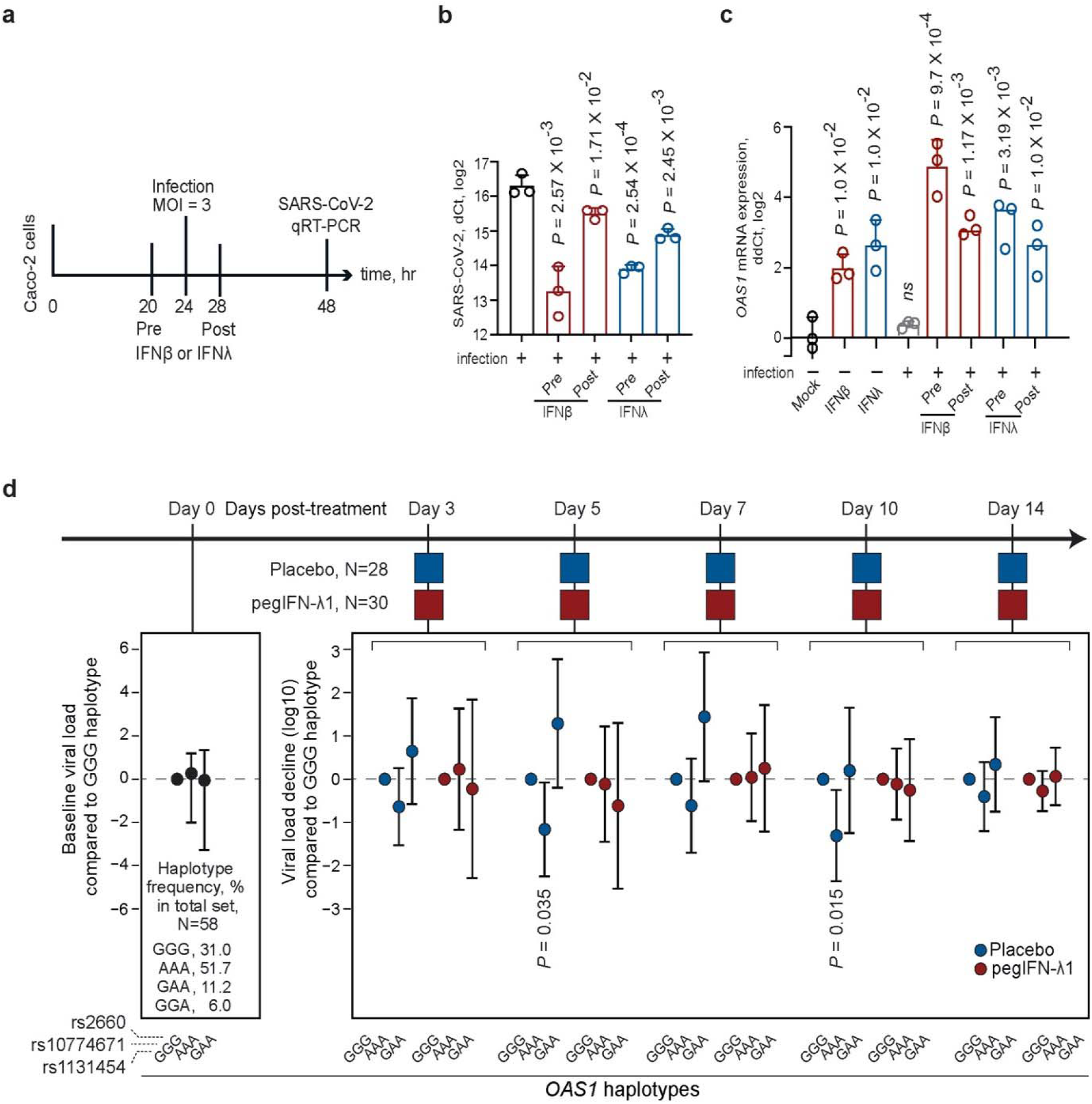
Effects of interferons on SARS-CoV-2 viral loads *in vitro* and in patients in a clinical trial. **a)** Outline of an experiment in Caco2 cells: cells were infected with SARS-CoV-2 and treated with IFNβ or IFNλ four hours before or after infection, and SARS-CoV-2 and *OAS1* expression was measured by qRT-PCR 24 hrs post-infection. Expression of **b)** SARS-CoV-2 and **c)** *OAS1* normalized by the expression of endogenous control (*HPRT1*). P-values are for comparison with infection alone or Mock; **d)** Outline of the clinical trial: a single subcutaneous injection of 180ug pegIFNλ1 or saline placebo was administered at day 0 and decline of SARS-CoV-2 load (log10 copies per ml) was evaluated at indicated days compared to day 0. The viral load decline is presented comparing to OAS1-GGG haplotype (rs1131454-G, rs10774671-G, rs2660-G) and adjusting for age, sex, and viral load at day 0 (complete results are shown in **Table S12**).

To investigate whether this can be relevant in clinical settings, we analyzed data from a clinical trial in which patients with mild outpatient COVID-19 were treated with a single subcutaneous injection of pegIFN-λ1 or saline placebo^18^. We genotyped OAS1 variants rs1131454, rs1074671, and rs2660 (capturing main *OAS1* haplotypes associated with COVID-19 outcomes in our case-case analyses) in genomic DNA of 58 participants. In multivariable analyses, baseline viral load was not associated with age, sex, race, treatment group, or *OAS1* haplotypes. Only baseline viral load and treatment group (placebo/treatment) were significantly associated with the viral decline in longitudinal analyses (**Table S12**). In the placebo group, which represents spontaneous viral clearance, compared to the GGG haplotype (*OAS1-p46*, Neandertal, non-risk), the association of AAA haplotype (*OAS1-p42*-A, risk) with an impaired viral loss showed similar trends at several days, reaching significance at day 5 and 10 post-treatment (**Figure 7d, Table S12**). However, the viral loss was not associated with *OAS1* haplotypes in the treatment group, indicating that pegIFN-λ1 treatment overcame the deficit in *OAS1* expression associated with the AAA haplotype (**Figure 7d, Table 12**). Thus, the *OAS1*-AAA haplotype was associated with both increased risk of hospitalized COVID-19 and impaired spontaneous clearance of SARS-CoV-2.

## DISCUSSION

Interindividual variability in response to SARS-CoV-2 infection resulting in the range from asymptomatic to fatal disease requires an urgent understanding of the underlying molecular mechanisms. Results of our cases-case analyses in individuals of European ancestry confirmed the genetic association within chr12q24.13 locus initially reported comparing patients with COVID-19 to the general population^2^. We fine-mapped the risk of hospitalized disease to a 19Kb-haplotype that included 76 *OAS1* variants forming a 95% credible set for association in this region. In functional analyses, we showed that alleles of two *OAS1* variants - a splicing variant rs10774671 and a missense variant rs1131454 in exon 3 regulate expression levels of *OAS1* through NMD. We demonstrated that the *OAS1* haplotype most targeted by NMD was associated with the lowest baseline *OAS1* expression, increased risk of severe COVID-19, and impaired spontaneous clearance of SARS-CoV-2, which could be compensated by early treatment with pegIFN-λ1.

A 185Kb-haplotype was reported as introgressed into the genomes of modern humans from the Neandertal and Denisova lineages of non-African ancestors^5-7,20^. It was hypothesized that the introgression of this haplotype was beneficial, perhaps by protecting humans from select pathogens; several associations with immune-related phenotypes reported for individual variants comprising this haplotype^8-16^ supported this idea. However, all these studies explored only one or a few variants, not providing sufficient clarity on the extent of the protective part of the Neandertal haplotype or the plausible molecular mechanisms underlying its protective effect. The analysis of COVID-19-related outcomes affords a unique opportunity to address these questions. Based on the COVID-19 GWAS meta-analysis, a 97Kb block of linked variants (r^2^>0.8) associated with hospitalized disease compared to the general population was nominated as a potentially protective part of the Neandertal haplotype^20^. Based on association and credible set analyses, we further narrowed this fragment to a 19Kb-region, which included 76 *OAS1* variants in r^2^>0.9 with the lead variant rs10774671 and comparably associated with the hospitalized disease.

Because rs10774671-G allele, associated with protection from severe COVID-19^1,2^, creates the OAS1-p46 protein isoform (vs. OAS1-p42 and some other isoforms), this marker can be considered the lead functional variant in the *OAS1* credible set^8,9^. Previous reports^21^ and our results showed that OAS1-p46 expression is enriched in trans-Golgi compartments, while OAS1-p42 is expressed in the cytosol. Targeting OAS1 to endomembrane structures may benefit response to pathogens that hide from cytosolic pattern recognition receptors^31^. Virus-induced formation of complex membrane rearrangements originating from the endoplasmic reticulum was demonstrated as the mechanism used by plus-strand RNA flaviviruses (such as West Nile, hepatitis C, Dengue, Yellow Fever, and Zika viruses) to evade sensing and elimination^31^. Trans-Golgi localization of OAS1-p46 would support enhanced sensing and clearance of flaviviruses, contributing to the strong differences in antiviral activities observed between OAS1-p42 and p46 isoforms^21^.

It was hypothesized that coronaviruses and SARS-CoV-2 specifically might use a similar mechanism to evade the immune response^32^. In embryonic kidney cells stably expressing ACE2 (HEK293-ACE2), SARS-CoV-2 was cleared in the presence of both OAS1 protein isoforms but more efficiently in the presence of OAS1-p46 than OAS1-p42^21^. We tested the functional effects of rs10774671 and several linked missense *OAS1* variants by creating corresponding *OAS1* plasmids. In A549-ACE2 cells used as our experimental model, all *OAS1* plasmids provided a similar anti-SARS-CoV-2 response. The presence of the C-terminal OAS1-p46 CaaX motif, which is responsible for trans-Golgi targeting, was insufficient to explain differences in antiviral activities of OAS1-p42 and OAS1-p46^21^. Furthermore, OAS1-p42 and OAS1-p46 were similarly functional as enzymes activating the antiviral RNaseL pathway^21^. These protein isoforms overexpressed in A549 cells similarly blocked *in-vitro* replication of Dengue virus via an RNaseL-dependent mechanism^33^. Overall, previous and our results suggest that both OAS1-p42 and OAS1-p46 have anti-SARS-CoV-2 activity, which is likely to be context-dependent, such as determined by the requirement of localization to specific cellular compartments for viral sensing, and expression levels of each isoform.

Our expression analyses showed that in all datasets explored, mRNA expression of OAS1-p46 transcript was on average 3.9-fold higher than *OAS1-p42*. We did not find evidence for transcriptional regulation of *OAS1* expression but demonstrated that *OAS1* expression is regulated by NMD differentially affecting *OAS1* isoforms. By creating an NMD-resistant *OAS1*-p46 transcript, the non-risk rs10774671-G allele plays the major role in preserving the *OAS1* expression. Additionally, we identified rs1131454 within exon 3 of *OAS1* as the second variant contributing to the regulation of *OAS1* NMD. Although rs1131454 is a missense variant in exon 3 (Gly162Ser), it did not appear to have a functional impact on the enzymatic activity of recombinant OAS1 proteins^34^ and in our anti-SARS-CoV-2 assays. Instead, the non-risk rs1131454-G allele of this variant creates an ESE/ESS to include the Short vs. Long forms of exon 3. Although NMD targets all *OAS1-p42* transcripts, transcripts with rs1131454-G allele are partially rescued from NMD. Because the *OAS1-p46* transcripts always carry the rs1131454-G allele, they are most NMD-resistant. Trans-Golgi accumulation of OAS1-p46 coupled with its increased expression due to the combined effects of non-risk rs10774671-G and rs1131454-G alleles may significantly enhance the antiviral activity of OAS1, offering a functional mechanism underlying the association of the Neandertal haplotype with COVID-19 outcomes.

We observed a decrease in SARS-CoV-2 expression after treating cells with interferons (either IFNβ or IFN-λ cocktail) before or after infection, suggesting that interferons can overcome insufficient viral clearance. Indeed, in a clinical trial with pegIFN-λ1, we observed that spontaneous but not the interferon-induced clearance of SARS-CoV-2 was associated with OAS1 haplotypes. Thus, our results suggest that spontaneous clearance of SARS-CoV-2 impaired in the presence of specific *OAS1* variants can be compensated for by treatment with interferons. Although this treatment accelerated viral clearance in all patients, patients with the risk *OAS1* haplotype (AAA for rs1131454-A, rs10774671-A, and rs2660-A) would benefit from this treatment the most because of their impaired ability to clear the virus without treatment. In our clinical trial, patients were treated with a single subcutaneous injection of pegIFN-λ1^18^. Due to the restricted expression of receptors, IFN-λ1, a type III interferon, is well tolerated without causing systemic side effects often associated with administration of type I interferons^35^. Inhaled nebulized type I interferons, IFNβ-1a^36^ and IFN-α2b^37,38^, are also tested as an early treatment for SARS-CoV-2 infection, with promising results.

The strengths of our study include genetic analyses evaluating outcomes in individuals with laboratory-confirmed COVID-19 and integrated analyses of multiple genomic datasets (e.g., RNA-seq, ATAC-seq, ChIP-seq, and Hi-C) supported by experimental testing of our hypotheses. In addition, we analyzed multi-variant haplotypes in association studies with COVID-19 severity and a clinical trial with pegIFN-λ1. The extensive multi-method investigation provides strong plausibility for our findings. Of multiple associated genetic variants, we identified rs10774671 and rs1131454 as most functional within *OAS1* for clearance of SARS-CoV-2 and COVID-19 severity. However, we cannot exclude additional functional variants, especially in non-European populations, in which we had low statistical power for genetic analyses. Some of the associations with severe COVID-19 might be related to the function of OAS3, which we did not explore. Specifically, in Europeans, we identified two missense *OAS3* variants that might be independently contributing to the risk of severe disease. The activity of OAS1 and OAS3 enzymes might be synergistic but cell type and condition-dependent, which further genetic and functional studies should explore. Our study did not explore the genetics of immune response to SARS-CoV-2 variants.

Overall, we propose that non-risk alleles of two variants (rs10774671-G and rs1131454-G) protect *OAS1* transcripts from NMD (**Figure 8**). The rs10774671-G allele has the major role by generating the *OAS1*-p46 isoform, while rs1131454-G additionally and independently contributes by creating an ESE that increases inclusion of Long exon 3, thus protecting both *OAS1-p46* and *OAS1-p42* from elimination by NMD. The non-risk G alleles of both variants create the most abundant and NMD-resistant *OAS1* isoform (*OAS1*-*p46*-Long). In contrast, the risk A alleles of both variants create the NMD-vulnerable and low-expressed isoform (*OAS1*-*p42*), while the haplotype with rs10774671-A but rs1131454-G allele creates the OAS1-p42 isoform with an intermediate NMD resistance and expression levels (**Figure 8**). Deficient spontaneous clearance in individuals carrying these risk OAS1 haplotypes can be compensated by early treatment with interferons, which should be further explored in clinical trials.

**Figure 8.**
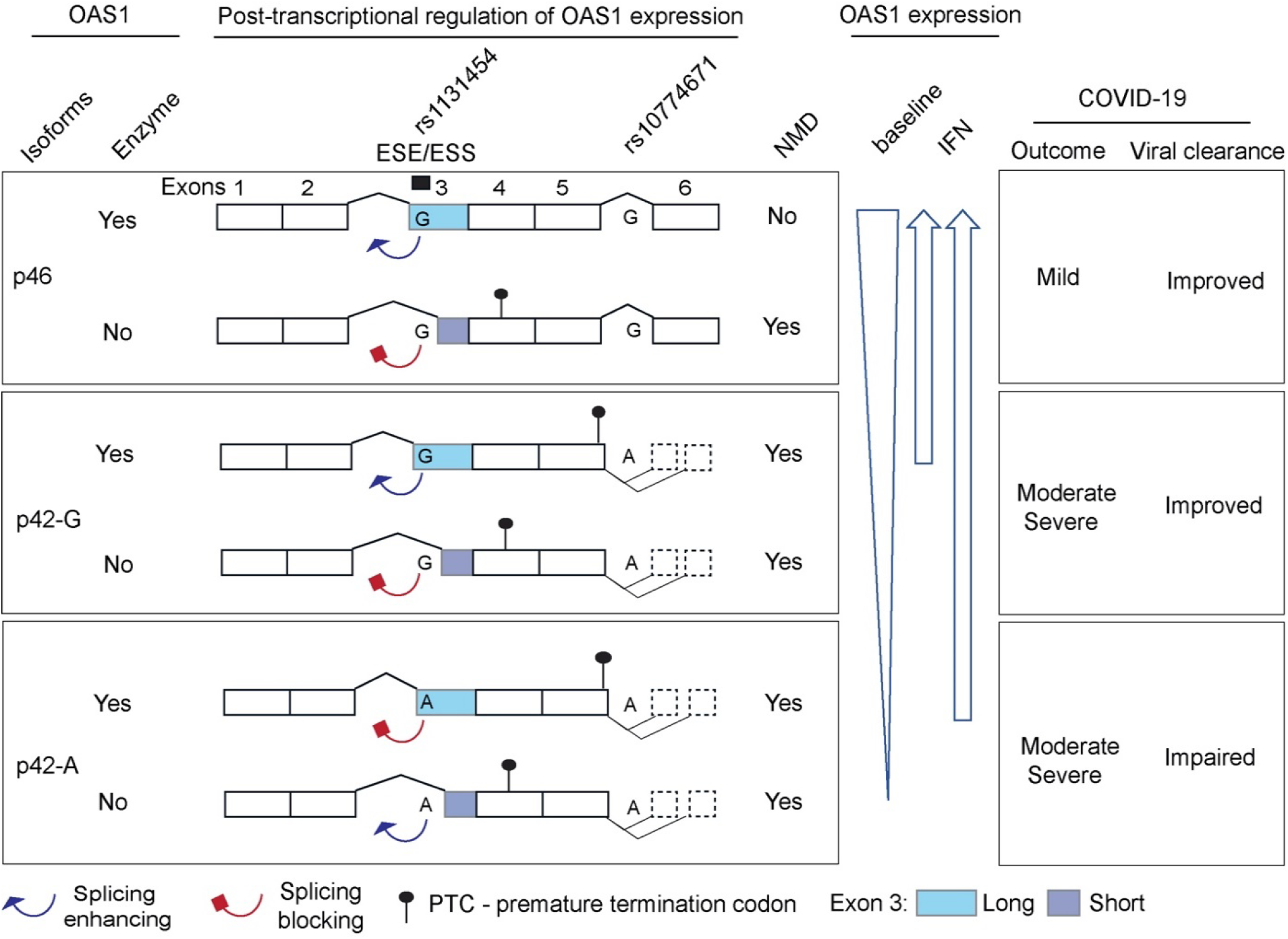
The proposed model for mechanisms underlying association between *OAS1* genetic variants and COVID-19 outcomes. Two *OAS1* variants – exon 3 missense variant (rs1131454-A/G, Gly162Ser) and splice site variant (rs10774671-A/G) determine the structure and expression levels of OAS1 isoforms. Alleles of the splicing variant rs10774671 define the *OAS1* isoforms – *OAS1-p42* (A allele) and *OAS1-p46* (G allele). Alleles of rs1131454 create an exonic splicing enhancer/silencer (ESE/ESS) for splicing of the canonical/Long vs. alternative/Short exon 3. Transcripts with Short exon 3 are terminated by premature stop codons (PTCs) within exon 4 and efficiently targeted by nonsense-mediated decay (NMD). The stop codon for *OAS1-p42* is located within exon 5, followed by several additional exons creating *OAS1-p44* and *OAS1-p48* isoforms, making this stop codon a PTC. Thus, *OAS1-p42* is also targeted by NMD, albeit less efficiently compared to transcripts with PTCs in exon 4. The combined splicing effects of rs10774671, which creates alternative *OAS1* isoforms and rs1131454, which regulates the inclusion of Short or Long exon 3 and thus the introduction of additional PTCs result in variable degradation of *OAS1* transcripts by NMD. At baseline, the expression is highest for *OAS1-p46* and the lowest for *OAS1-p42*-A. However, treatment with IFNs may compensate for NMD, allowing OAS1-p42 protein to reach expression levels comparable to OAS1-p46. Thus, the effects of genetic variants on OAS1 expression can be compensated by IFN treatment to overcome impaired viral clearance leading to severe COVID-19.

## Data Availability

Summary statistics for all genetic analyses is provided in Supplementary Tables. Individual genotypes will be submitted to dbGAP. Dataset for Oxford Nanopore RNA-seq was deposited to NCI SRA as BioProject PRJNA743928. Full-length sequence data for OAS1-p42 transcript with Short exon 3 was deposited to NCBI GenBank with accession number MZ491787. Requests for any additional data or reagents should be addressed to L.P.-O.

## ACKNOWLEDGEMENTS

We are grateful to all the patients who donated their samples. We are also indebted to all the doctors and nurses who contributed to this study. Additionally, we thank Dr. Lisa McReynolds (CGB/DCEG/NCI) for clinical review of the samples, Dr. Mitchell Machiela (ITEB/DCEG/NCI) for contributing to data analysis plan, Melinda Yan (LTG/DCEG/NCI) for help with analysis, Nathan Cole (CGR/DCEG/NCI) for help with the acquisition of TCGA sliced BAM files, Dr. Michael Dean (LTG/DCEG/NCI) for help with Oxford Nanopore sequencing, Giannis Vatsellas and Artemis G. Korovesi (Biomedical Research Foundation of the Academy of Athens) for technical assistance on DNA extraction and preparation, Julie C Sapp (NHGRI) for support and guidance in patient recruitment, Maureen Martin (BSP, Frederick National Laboratory for Cancer Research) for coordinating sample and data transfers, Jonathan Li (Harvard Medical School) for cohort organization, Lianhua Jin, Jing Wu and Rachael Shevin in the UAB Center for Clinical and Translational Science for preparation of genomic DNA. The results are partially based on data generated by the TCGA Research Network. We used data provided by the Kaiser Permanente Research Bank (KPRB) from the KPRB collection, which includes the Kaiser Permanente Research Program on Genes, Environment, and Health (RPGEH), funded by the National Institutes of Health (RC2 AG036607), the Robert Wood Johnson Foundation, the Wayne and Gladys Valley Foundation, The Ellison Medical Foundation, and the Kaiser Permanente Community Benefits Program. Access to data used in this study may be obtained by application to the KPRB at kp.org/researchbank/researchers. Research reported in this publication was supported by the Intramural Research Programs of the National Cancer Institute: Division of Cancer Epidemiology and Genetics and Center for Cancer Research, Frederick National Laboratory for Cancer Research, contract number HHSN261200800001E; National Center for Advancing Translational Science of the National Institutes of Health under award number UL1TR003096 associated with the University of Alabama at Birmingham COVID-19 Enterprise Study IRB-20005127, NHGRI Intramural Research Program grants HG200388-07 and HG200359-12. Additional funding was provided by the state of Alabama through the Alabama Genomic Health Initiative (IRB F170303004). MLS was supported by grants: DFG (416072091) and the BMBF (01KI20239B). SB was supported by Deutsche Forschungsgemeinschaft (DFG): project numbers 415089553 (Heisenberg program), 240245660 (SFB1129), and 272983813 (TRR179), the state of Baden Wuerttemberg (AZ: 33.7533.-6-21/5/1) and the Bundesministerium Bildung und Forschung (BMBF) (01KI20198A). EA was supported by research grants from the European Commission – IMMUNAID (779295) and CURE (767015) and the Hellenic Foundation for Research and Innovation INTERFLU (1574). HBK was supported by grant no 02-2020-012 from the SNUBH Research Fund. The pegIFN-λ1 clinical trial was supported by the Toronto COVID-19 Action Initiative (72059280), the Ontario Together COVID19 Research Application (C-224-2428560-FELD) and the Canadian Institutes for Health Research (VR3-172648). Support for title page creation and format was provided by AuthorArranger (https://authorarranger.nci.nih.gov/), a tool developed at the NCI. The content of this publication is solely the responsibility of the authors and does not necessarily represent the official views and policies of the National Institutes of Health or Department of Health and Human Services, nor does mention of trade names, commercial products, or organizations imply endorsement by the U.S. Government.

## Author Contributions

A.R.B. and L.P.-O. conceived and designed the study. L.P.-O. supervised the study, A.R.B., M.L.S., O.O.O., S.B., J.F. and L.P.-O. designed the experiments and analyses. A.R.B., M.L.S., O.O.O., M.A.Z., B.W.P., T.J.R., M.H. and J.V. performed the experiments and analyzed the data. O. F.-V. analyzed genetic data, H.P.S.A and M.Y. contributed to the analysis of genetic data. A.R.B. and C.-H.L. performed computational analyses of genomic data. S.J.C. supervised the COVNET study. A.A.H. supervised COVNET sample processing and genotyping. L.M., S.S., and V. V. contributed to COVNET sample and data management. A.R.B., M.L.S., O.F.V., O.O.O., M.A.Z., E.A., G.B., L.G.B., D.L.B., A.B.-H., M.C., E.C., P.G.C., R.L,C., C.D., J.E., N.E., H.S..F., G.S.F., A.J.G., S.H., A.A.H., H.I., M.G..I., H.B.K., R.J.K., B.R.K., J.A.P., M., J.P., D.J.R., D.T.R., M.D. R., B.R., E.S., V., T., A.V., D.R.W., X.Y., Y.Z., S.B., S.J.C., J.F., L.P-O. contributed reagents/materials/analysis tools. A.R.B. and L.P.-O. wrote the manuscript. All authors discussed the results and commented on the manuscript.

## METHODS

### Patients from COVNET

Patients were recruited by several studies participating in COVNET: Large-scale Genome-wide Association Study and Whole Genome Sequencing of COVID-19 Severity (https://dceg.cancer.gov/research/how-we-study/genomic-studies/covnet), based on ethical approvals by corresponding institutions and informed consents provided by patients. COVID-19 diagnosis was confirmed based on positive viral testing or serology. Sample collection occurred pre-emegence of SARS-CoV-2 variants. Detailed demographic and clinical records were provided by the participating studies and independently reviewed by COVNET team. Outcomes were defined as mild (non-hospitalized) COVID-19, hospitalized moderate (not requiring mechanical ventilation) and hospitalized severe (mechanical ventilation or death due to COVID-19).

We used data and samples from a clinical trial (NCT04354259), in which patients with mild outpatient COVID-19 received a single subcutaneous injection of 180 µg pegIFN-λ1 or saline placebo^18^. DNA was extracted from PBMCs of all participants and genotyped for 3 *OAS1* variants – rs1131454, rs10774671, and rs2660 representing main risk and non-risk haplotypes. SARS-CoV-2 RNA decline (viral copies, log10) at baseline and several days after treatment were analyzed in relation to *OAS1* haplotypes, adjusting for relevant covariates.

### Analysis of COVNET data

COVNET DNA samples were analyzed by AmpFLSTR Identifiler and then genotyped for 712,191 variants using the Global Screening Array version 2.0 (GSA2, Illumina) by the Cancer Genomics Research (CGR) Laboratory, DCEG/NCI. Samples exceeding an estimated contamination rate > 20% by VerifyIDintensity, an autosomal heterozygosity deviation FHET within +/- 0.20, a missing call rate >2%, and sex-discordant samples were excluded. Variants deviating from Hardy– Weinberg equilibrium (p < 10e-6), with a minor allele frequency < 0.01, an overall genotyping rate < 95%, or differential missingness between cases and controls (p-value cutoff 0.001) were excluded. Additionally, whole-genome sequencing data generated by contributing studies were used for a subset of samples. Sex, relatedness and population groups (European, African, admixed-Hispanic and East-Asian) were determined based on genetic analyses for all patients. All genotyped variants were used for principal components analysis (PCA) with GCTA (v1.93.0 beta) to generate the first 20 eigenvectors, separately for each ancestry.

Imputation of additional variants was performed using the Michigan Imputation Server with the Minimac4 algorithm. We imputed the whole chromosome 12 based on population-specific reference panels: the Trans-Omics for Precision Medicine (TOPMed) reference panel was used for individuals of European ancestry; the Consortium on Asthma among African-ancestry populations in the Americas (CAAPA) reference panel was used for individuals of African ancestry; the American admixture population from Phase 3 of the 1000 Genomes Project (AMR-1KGP) reference panel was used for individuals of Hispanic-ancestry; and the Genome Asia pilot (GAsP) reference panel was used for individuals of East Asian-ancestry. Imputed variants with R^2^ > 0.9 were retained for further analyses. SNPs rs10774671, rs1131454, and rs2285933 were directly genotyped with TaqMan assays in all non-European populations due to low imputation quality for these variants and in a subset of European individuals to confirm high concordance with imputed genotypes.

Each variant coded as 0, 1, or 2 based on the counts of the variant allele was analyzed for association using logistic regression with glm function with binomial family and logit link in R software (v4.0.4). For haplotype analyses, ShapeIT (v2.r837) software was used to phase the selected variants using the genetic map file for chromosome 12 with recombination frequencies for GRCh38/hg38. Haplotype association analyses using logistic regression were done with PLINK (v1.07), with a custom selection of a reference haplotype. Both single-variant and haplotype analyses were performed adjusting for sex, age, squared mean-centered age, and the first 20 principal components as covariates to control for population stratification. Summary statistics for association analyses for hospitalized vs. non-hospitalized patients of European ancestry were used to generated 95% credible sets using Sum of Single Effects (SuSiE) R package (v0.9.26)^19^, where the R2 LD matrix was generated from the genotype data using the genetics R package (v1.3.8.1.3). LD plots were generated with Haploview 4.2.

### Cell lines

Details for all cell lines used in this work are presented in **Table S13**. Cell lines were either used within 6 months after purchase or periodically authenticated by microsatellite fingerprinting (AmpFLSTR Identifiler, ThermoFisher) by the Cancer Genomics Research Laboratory/DCEG/NCI. All cell lines were regularly tested for mycoplasma contamination using the MycoAlert Mycoplasma Detection kit (Lonza).

### TaqMan genotyping

Genotyping of rs10774671, rs1131454, rs2660, and rs2285933 was done using TaqMan genotyping assays (**Supplementary Table S14**), with 2x TaqMan expression Master Mix (Qiagen) and 2-5 ng of genomic DNA in the 5-μl reactions in 384-well plates on QuantStudio 7 (ThermoFisher). Positive controls (HapMap samples with known genotypes) and negative controls (water) were included on each 384-well plate.

### Plasmids

Plasmids with a FLAG-tag for *OAS1-p46*-G (ID: OHu21619D, rs1131454-G, rs1131476-G, rs1051042-G) and *OAS1-p42*-A (ID: OHu29197D, rs1131454-A) were purchased from GenScript. QuikChange II site-directed mutagenesis kit (Agilent 200523) was used to generate plasmids *OAS1-p42*-G (rs1131454-G) and *OAS1-p46*-A (rs1131454-A, rs1131476-A, rs1051042-C) using mutagenesis primers (**Table S14**). Sequences of original and modified plasmids were confirmed by Sanger sequencing.

### SARS-CoV-2 infections

SARS-CoV-2 (strain BavPat1) was obtained from the European Virology Archive, amplified in Vero E6 cells, and used at passage 3. Media was removed from plated cells, and SARS-CoV-2 (MOI 3) was added to cells for 1 hour at 37°C; then the virus was removed, cells were washed 1x with PBS, and fresh media was added back to the cells. RNA from harvested cells was extracted using RNeasy kit (Qiagen), cDNA was generated with iSCRIPT reverse transcriptase (BioRad) from 250 ng of total RNA, and qRT-PCR was performed using SYBR Green assays (iTaq SYBR Green buffer, BioRad) or TaqMan expression assays (**Table S14**) and as previously described^29,30^.

A549-ACE2 cells (2.0×10^5^ cells) were seeded in 12-well plates 24 hrs before transfection. Cells were transfected with the indicated *GFP* or *OAS1* plasmids using Lipofectamine 2000. Media was replaced 6 hrs post-transfection, and cells were infected with SARS-CoV-2 at an MOI=3 for 1 hr 48 hrs post-transfection. Following infection, fresh media was added, and SARS-CoV-2 replication was monitored for 24 hrs post-infection.

Caco2 cells (7.5×10^4^) were seeded in 48-well plates for 20 hrs, then media was removed, and interferons were added to the wells for 4 hrs. Media with interferons was collected and added back after infection with SARS-CoV-2 for 1 hr. Alternatively, Caco2 cells were infected with SARS-CoV-2 for 1hr first and then 4 hrs post-infection media with the virus was replaced with media with interferons for the indicated time. Interferon treatment: 2000 IU/mL of IFNβ or 300 ng/mL (a cocktail of 100ng each of IFN-λ1, IFN-λ2 and IFN-λ3).

### Confocal microscopy

HT1376 bladder cancer cell line (rs10774671-GG genotype, OAS1-p46 isoform) and A549 lung cancer cell line (rs10774671-AA genotype, OAS1-p42 isoform) were plated on 4-well chambered slides (2×10^4^ cells per well, LabTek) for 24 hrs. Cells were left untreated or treated with 2ng/ml IFNβ (R&D System) for 24 hrs. Cells were then washed twice with PBS and fixed with 4% paraformaldehyde (BD Biosciences) for 30⍰min. After rinsing twice in PBS and permeabilization buffer (BD Biosciences), cells were incubated with permeabilization buffer for 1⍰hr. Fixed cells were incubated with mouse anti-Golgin-97 antibody (1:250 dilution, ThermoFisher, A-21270) for 3 hrs at room temperature, washed, and then stained with anti-rabbit Alexa Fluor 488 (1:500 dilution, ThermoFisher, A21202). Cells were then incubated with rabbit anti-OAS1 antibody (1:100 dilution, ThermoFisher, PA5-82113) overnight, washed, and stained with anti-rabbit Alexa Fluor 680 (1:500 dilution, ThermoFisher, A10043). Slides were mounted with antifade mounting media with DAPI (Thermo Fisher) and imaged at 63x magnification on an LSM700 confocal laser scanning microscope (Carl Zeiss) using an inverted oil lens. Colocalization and correlation coefficients between OAS1 and Golgin-97 expression were generated with LSM700 Zen software by analyzing randomly imaged fields of view (5-7 fields) containing at least 7 cells from IFNβ-treated wells. The linear relationship between the expression of OAS1 and Golgin-97 at every pixel with protein expression was determined with Pearson’s correlation coefficient^39^. Cells with less than 10 analyzed pixels were excluded due to very low expression of either protein, making the correlation data unreliable. Mander’s overlap coefficients were also calculated^39^, which factor in the total number of pixels of either protein.

### RNA-seq analysis of data from NCBI SRA and TCGA

RNA-seq datasets were accessed from the NCBI sequence read archive (SRA) by using SRA command-line tools. SRA datasets analyzed in this study are listed in **Table S15**. Briefly, the raw FASTQ files were aligned with STAR version 7.1.31 to the reference human genome assembly (hg38). Low-quality sequencing files with ≤80% of mappable reads were excluded from further analyses. BAM slices were indexed and sliced to include 117 Kb of the *OAS1-3* genomic region, chr12:112,901,893-113,019,729, hg38. For TCGA, BAM slices for the *OAS* gene locus were generated through the NCI Genomics Data Commons portal accessed on 25 November 2020, using standard workflow (https://docs.gdc.cancer.gov/API/Users_Guide/BAM_Slicing/).

### Estimation of RNA-seq read counts specific to *OAS1* isoforms

Expression of *OAS1* isoforms - *p42, p44, p46*, and *p48* was quantified based on unique RNA-seq reads. Specifically, RNA-seq BAM slices were processed using the R package ASpli version 1.5.1 with default settings, and exon and exon-exon junction specific reads were quantified and exported in a tab file format. For *OAS1* isoforms - *p44, p46*, and *p48*, RNA-seq reads specific to their unique last exon-exon junctions were used for quantification. For the *p42* isoform, which does not have a unique exon-exon junction, sequencing reads corresponding to its unique 3’UTR (extension of exon 5) were used as a proxy for quantification. For normalizing the expression, junction reads were divided by 50 (average length of an RNA-seq read), and *p42* 3’ UTR exon reads were divided by 317 bp, corresponding to its length. The mean expression of each isoform was calculated from samples with ≥3 RNA-seq reads supporting the unique splice junction or exon.

### Analysis of ATAC-seq, ChIP-seq, and Hi-C data of cell lines

The ATAC-seq, H3K27ac ChIP-seq, Hi-C, and RNA-seq raw data for SW780, HT1376, and SCABER bladder cancer cell lines were downloaded from NCBI SRA (ID: PRJNA623018) using the SRA tools. For ATAC-seq and H3K27ac ChIP-seq analysis, the FASTQ files were aligned to hg19 using Encode-DCC ATAC-seq-pipeline (https://github.com/ENCODE-DCC/atac-seq-pipeline) and chip-seq-pipeline2 (https://github.com/ENCODE-DCC/chip-seq-pipeline2) with default settings. The output bigwig files were then uploaded to the UCSC genome browser for visualization. For RNA-seq analysis, the FASTQ files were mapped to hg19 using STAR aligner (https://github.com/alexdobin/STAR) with default settings. The output sorted BAM files were indexed using SAM tools (https://github.com/samtools/). For Hi-C, FASTQ files were processed using Juicer (https://github.com/aidenlab/juicer) by selecting relevant restriction cutting sites such as Mbo I/Dpn II and aligned to hg19. The chromatin loops in Hi-C data were detected using Hiccups in Juicer tools with default settings. The same procedure was applied to analyze Hi-C data for THP-1 monocytic cell line untreated or treated with INFb for 6 hrs. The Hi-C and chromatin interaction files were visualized in the UCSC genome browser (https://genome.ucsc.edu). Integrative data analysis was performed to identify open chromatin marks and chromatin interactions between potential genetic variant co-localizing with enhancers and promoters of OAS1, OAS2, and OAS3 within the COVID-19-associated genetic locus.

### Genotyping and allele-specific expression analysis from RNA-seq data

RNA-seq BAM slices were genotyped for OAS1 exonic variants with an IGV command-line tool using 21 bp sequence centered at each variant. For each variant, a 10% threshold of allele-specific reads was used for genotype calling.

### Analysis of exonic splicing enhancer activity affected by rs1131454 within *OAS1* exon 3

The allele-specific 27 bp sequence (GUCAGUUGACUGGC[A/G]GCUAUAAACUA) centered on rs1131454 was used for the prediction of exonic splicing enhancer (ESE)/silencer (ESS) motifs using the Human Splicing Finder (HSF, www.umd.be/HSF3/). The binding sites for alternative splicing factors were depicted with a bar graph. Exontrap mini-genes were generated for alleles of rs1131454. Specifically, allele-specific sequences of exon 3 with 100 bp of flanking intronic sequences and overhangs for restriction sites (Xho1 and Not1) were custom-synthesized as gene fragments (IDT, **Table S16**). These fragments were cloned in sense orientation in Exontrap vector pET01 (MoBiTec) using XhoI and NotI restriction sites and validated by Sanger sequencing. The A549 and T24 cells were seeded in a 12-well plate at a cell density of 2×10^5^ and transfected after 24 hrs with 200 ng of allele-specific mini-genes using Lipofectamine 3000 transfection reagent (Invitrogen) in 3 biological replicates. At 48 hrs post-transfection, cells were harvested, and total RNA was extracted with QIACube using RNeasy kit with on-column DNase I treatment (Qiagen). cDNA was prepared for each sample with 500 ng of total RNA using SuperScript III reverse transcriptase (Invitrogen) and a vector-specific primer: 5⍰-AGGGGTGGACAGGGTAGTG-3⍰. cDNA corresponding to 5 ng of RNA input was used for each RT-PCR reaction. Two common primer pairs were used for characterizing splicing products of allele-specific mini-genes (**Table S16**). Only primer pair 1 (FP vector exon 1: GGA GGA CCC ACA AGG TCA GTT; and RP exon 3: GCTG CTT CAG GAA GTC TCT CTG) identified alternative splicing events corresponding to endogenous exon 3 splicing between vector exon 1 and insert after PCR-amplified products were resolved by agarose gel electrophoresis. The specific bands were cut out from the gel, purified, and validated by Sanger sequencing. The ratio of alternative splicing products was calculated based on band intensity using densitometry, and fold changes were calculated between two allele-specific mini-genes.

### Oxford Nanopore RNA-seq

A549 or HT1376 cells (2×10^6^ per sample) were seeded in T25 flasks overnight. The next day media was replaced with either media containing 2 ng/mL IFNβ (treated) or normal media without IFNβ (mock). Total RNA was prepared from cells 24 hrs post-treatment using the RNeasy Mini Kit (Qiagen). PolyA+ RNA was enriched from total RNA using the Dynabeads mRNA Purification Kit (Invitrogen). cDNA libraries were prepared from 200 ng of PolyA+ RNA using the Direct cDNA Sequencing Kit (Oxford Nanopore), according to the PCR-free 1D read protocol for full-length cDNA (Oxford Nanopore, SQK-DCS109), with some modifications. Specifically, RNase Cocktail Enzyme Mix (ThermoFisher) was used during the RNA digestion step after the first-strand synthesis; all reaction amounts for reverse transcription reactions up to the second-strand synthesis step were doubled; from second-strand synthesis up to adapter ligation, reactions were 1.5x of original amounts; during adapter ligation, 35 ul Blunt/TA Ligase Master Mix was used instead of the recommended 50 ul, and nuclease-free water was excluded. Final libraries were loaded into MinION Fluidics Module flow cells (Oxford Nanopore, FLO-MIN106D), and sequencing was carried out on GridION MK1 and MinION MK1C instruments (Oxford Nanopore) for 3 days, using default parameters.

The FASTQ files generated by Nanopore GridION long-read sequencer were trimmed using Porechop (https://github.com/rrwick/Porechop) and aligned to the hg19 genome using Minimap2 (https://github.com/lh3/minimap2) with -ax splice command. The output SAM files were then converted to indexed, sorted BAM files using SAM tools (https://github.com/samtools/). The BAM files were visualized with the UCSC genome browser.

### Analysis of nonsense-mediated decay (NMD) of *OAS1* isoforms

We downloaded RNA-seq data for HeLa cells (OAS1-p42 expressing) with and without siRNA-mediated knockdown of NMD genes – *SMG6* and *SMG7* (SRA:PRJNA340370). The FASTQ files were aligned with STAR aligner (https://github.com/alexdobin/STAR) with default settings followed by quantification of isoforms-specific reads for alternative splicing junctions of *OAS1* exon 3 with adjacent exons. The data was also visualized as sashimi plots using Integrative Genome Viewer. We also generated a triple-knockdown (tri-KD) by transfecting A549 (OAS1-p42 expressing) and HT1376 (OAS1-p46 expressing) cells with siRNAs – scrambled (negative control) and targeting genes for the NMD pathway (*SMG6, SMG7*, and *UPF1*). After 48 hrs, cells were harvested, and total RNA was isolated using an RNeasy kit with on-column DNase I treatment (Qiagen). Subsequently, cDNA for each sample was prepared from equal amounts of RNA using the RT^2^ First-Strand cDNA kit (Qiagen). TaqMan assays were used to confirm the knockdown of each gene, using the expression of HPRT1 as an endogenous control (**Table S14**). Detection of *OAS1* exon 3 splicing events was done with custom expression assays (**Table S14**). For each condition, experiments were performed in biological triplicates, and expression was quantified in four technical replicates on QuantStudio 7 (Life Technologies) using TaqMan Gene Expression buffer (Life Technologies). Genomic DNA and water were used as negative controls for all assays. Expression was measured as C_t_ values (PCR cycle at detection threshold) and calculated as dCt values normalized by endogenous control and ddCt values normalized by a reference group of samples.

### Western blotting

Cells (Caco2, HT1376, A549, and HBEC) were plated in 6-well plates (5×10^5^ cells per well) and were untreated or treated with IFNβ (1ng/ml), IFNγ (2ng/ml) or IFN-λ3 (100ng/ml) for 24 hrs. Cells were lysed with RIPA buffer (Sigma) supplemented with protease inhibitor cocktail (Promega) and PhosSTOP (Roche) and placed on ice for 30⍰min, with vortexing every 10⍰min. Lysates were pulse-sonicated for 30⍰s, with 10⍰s burst-cooling cycles, at 4°C, boiled in reducing sample buffer for 5⍰min and resolved on 4–12% Bis-Tris Bolt gels and transferred using an iBlot 2 (ThermoFisher). Blots were blocked in 2.5% milk in 1% TBS-Tween before staining with rabbit anti-OAS1 antibody (1:200, Thermo Fisher, PA5-82113) and rabbit anti-GAPDH antibody (1:500, Abcam, ab9485). The signals were detected with HyGLO Quick Spray (Denville Scientific) or SuperSignal West Femto Maximum Sensitivity Substrate (ThermoFisher) and viewed on a ChemiDoc Touch Imager with Image Lab 5.2 software (Bio-Rad).

### Bioinformatic analyses

We utilized the computational resources of the NIH Biowulf supercomputing cluster (http://hpc.nih.gov) and specific packages for R version 3.6.2.

## Data availability

Summary statistics for all genetic analyses is provided in Supplementary Tables. Individual genotypes will be submitted to dbGAP. Dataset for Oxford Nanopore RNA-seq was deposited to

NCI SRA as BioProject PRJNA743928. Full-length sequence data for *OAS1-p42* transcript with Short exon 3 was deposited to NCBI GenBank with accession number MZ491787. Requests for any additional data or reagents should be addressed to L.P.-O.

## Supplementary Figures

**Figure S1.**
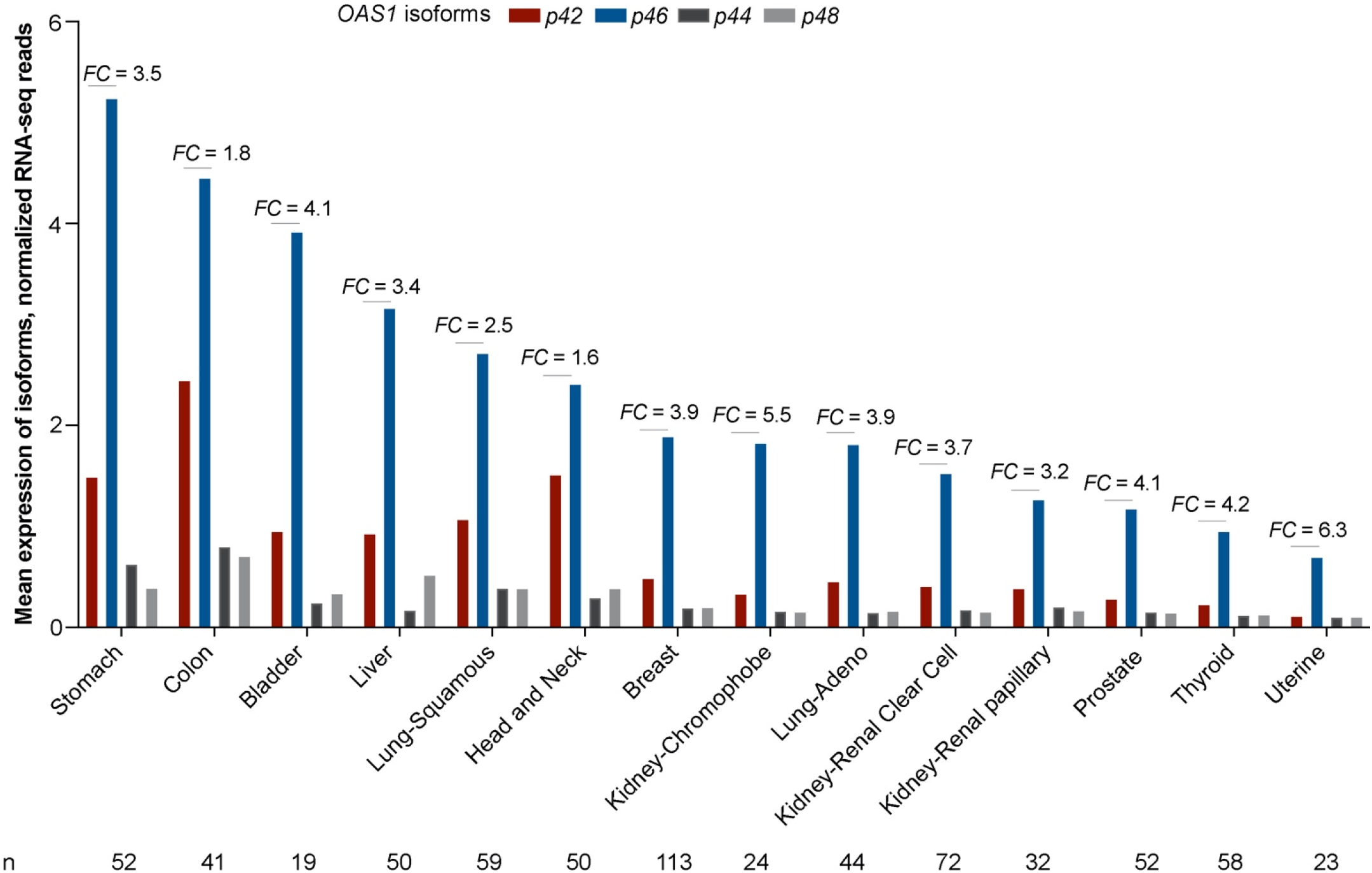
Mean expression levels of *OAS1* isoforms in adjacent normal tissues in TCGA. Expression of *OAS1* isoforms was analyzed in RNA-seq data in The Cancer Genome Atlas (TCGA), using tumor-adjacent normal tissues with RNA-seq data in ≥15 samples. RNA-seq reads for each *OAS1* isoform were calculated based on unique splice junctions (*OAS1-p44, p46*, and *p48*) or unique 3’UTR (*p42*). For normalization, the total number of RNA-seq reads for exon-exon junctions for *p44, p46* and *p48* was divided by 50 (length of an RNA-seq read) and unique RNA-seq reads for *p42* exon 5 by its length (317 bp). The mean expression levels of each isoform were calculated from all the samples with ≥3 RNA-seq reads. Overall, the mean expression of *OAS1-p46* is higher than *OAS1-p42*, with an average fold change (*FC*) of 3.9 across tissues.

**Figure S2.**
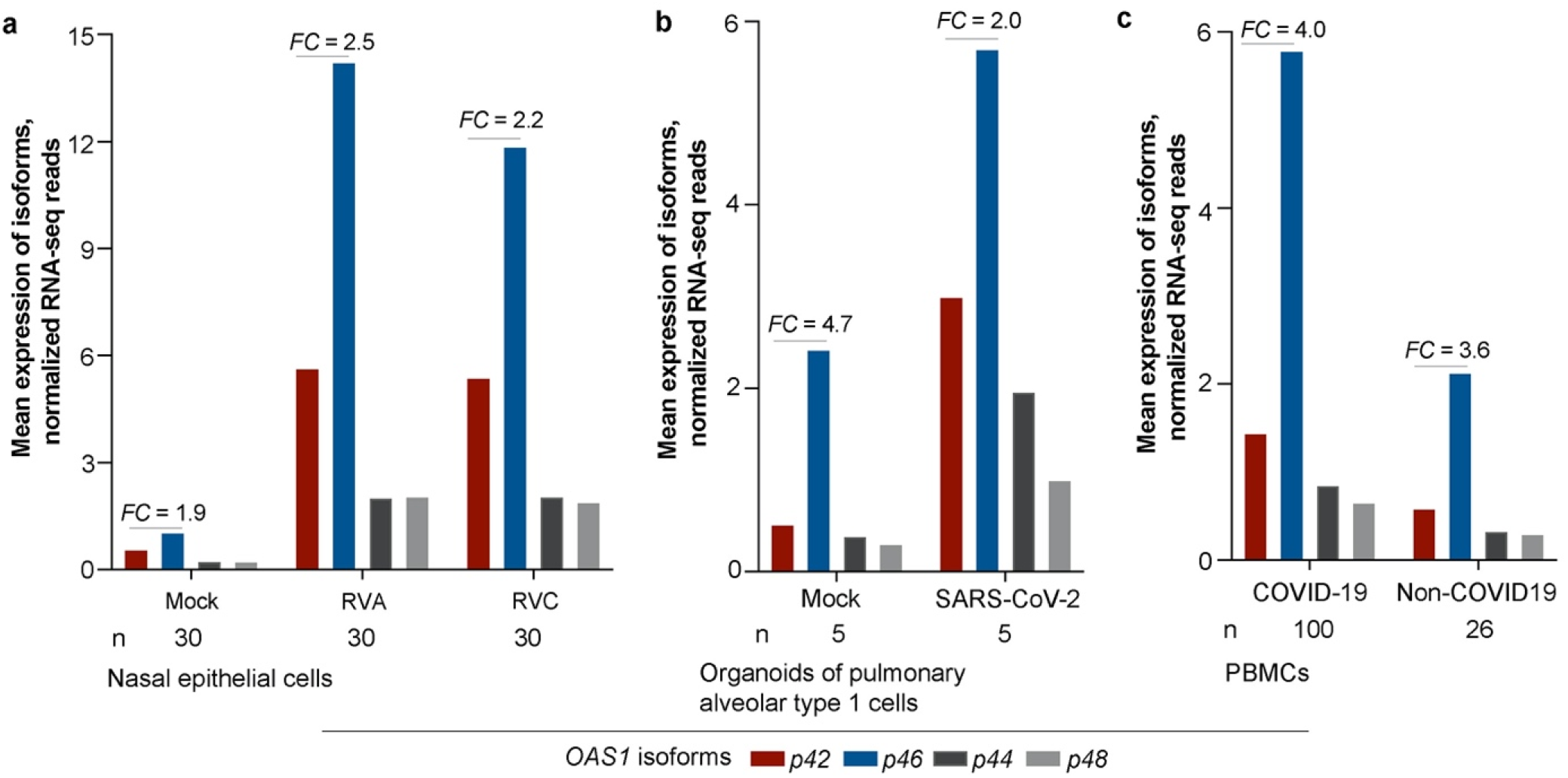
Mean expression levels of *OAS1* isoforms in nasal epithelial cells, pulmonary alveolar cells and PBMCs. Expression of *OAS1* isoforms was analyzed in RNA-seq data of **a)** nasal epithelial cells (SRA PRJNA627860), **b)** organoids of pulmonary alveolar type 1 cells (SRA PRJNA673197), and **c)** PBMCs (SRA PRJNA660067). The bar graphs show the mean expression levels of each *OAS1* isoform normalized as described in **Figure S1**. The expression of *OAS1-p46* is higher than *OAS1*-*p42* in all cell types and conditions tested.

**Figure S3.**
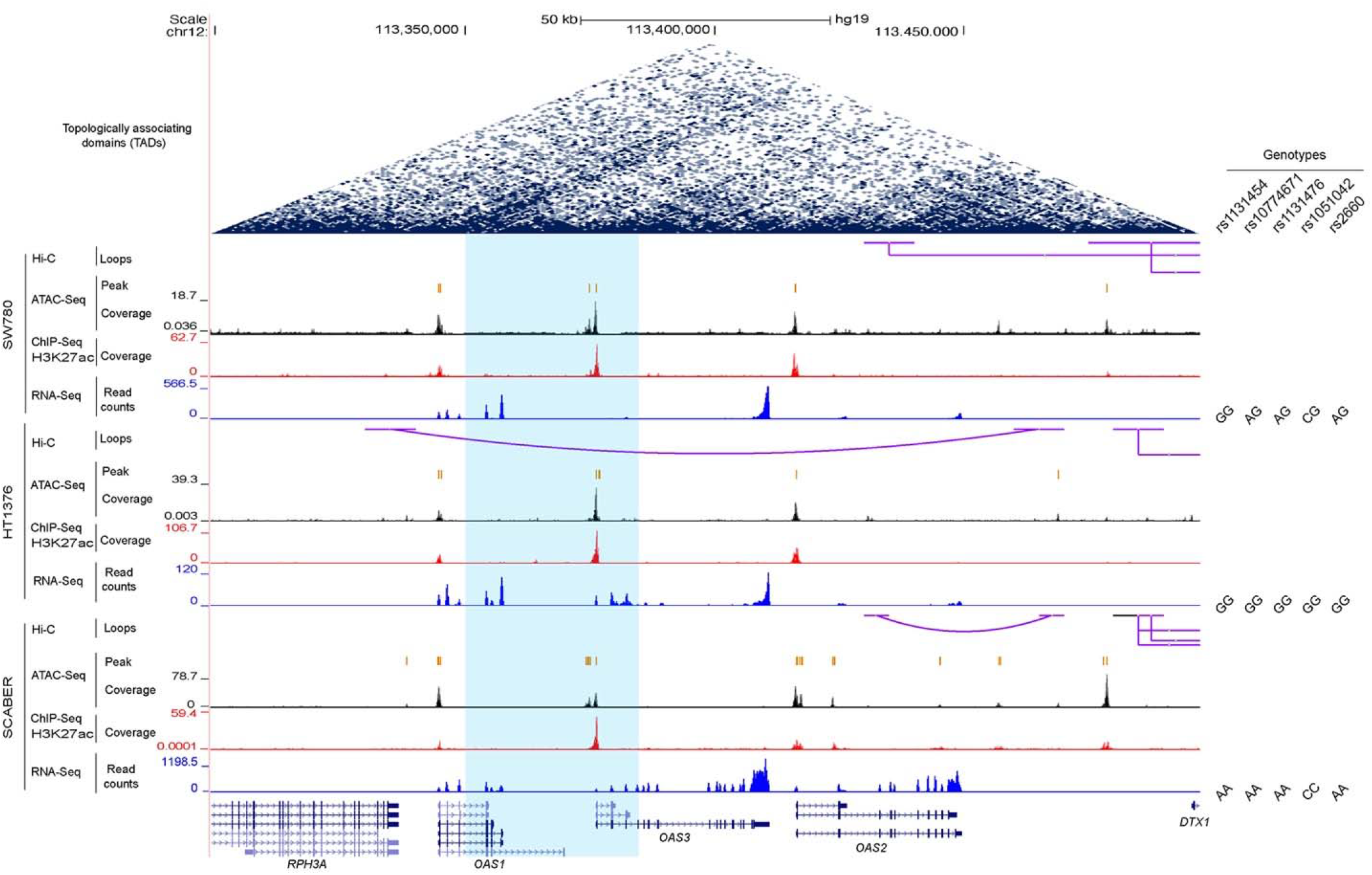
Multi-omics profile of the *OAS1*/*OAS3*/*OAS2* genomic region. Multi-omics data for genome-wide Hi-C, ATAC-seq, H3K27ac ChIP-seq, and RNA-seq of three bladder cancer cell lines (SRA PRJNA623018) were analyzed and visualized using UCSC genome browser. *OAS1, OAS2*, and *OAS3* expression was observed in all samples, but there is no evidence of open chromatin, enhancer activity, or chromatin interactions within the LD block (blue shading) that includes top 125 variants associated with hospitalized vs. non-hospitalized COVID-19 in Europeans **(Figure 1)**.

**Figure S4.**
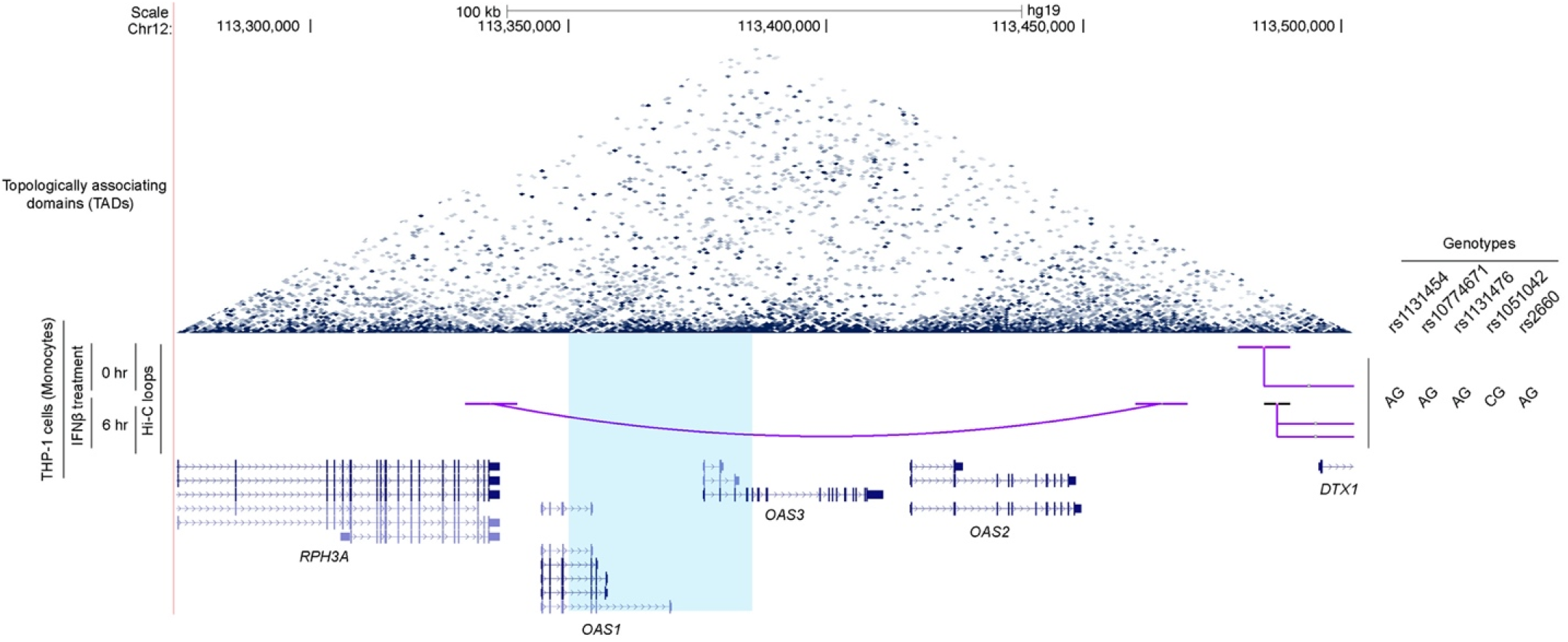
Hi-C chromatin interaction analysis in the *OAS1/OAS3/OAS2* genomic region. Chromatin interaction (Hi-C) data for the THP-1 monocytic cell line untreated and treated with IFNβ for 6 hrs (SRA PRJNA401748) were analyzed and visualized using UCSC genome browser. No chromatin interactions were detected with the LD block (blue shading) that includes top 125 variants associated with hospitalized vs. non-hospitalized COVID-19 in Europeans **(Figure 1)**.

**Figure S5.**
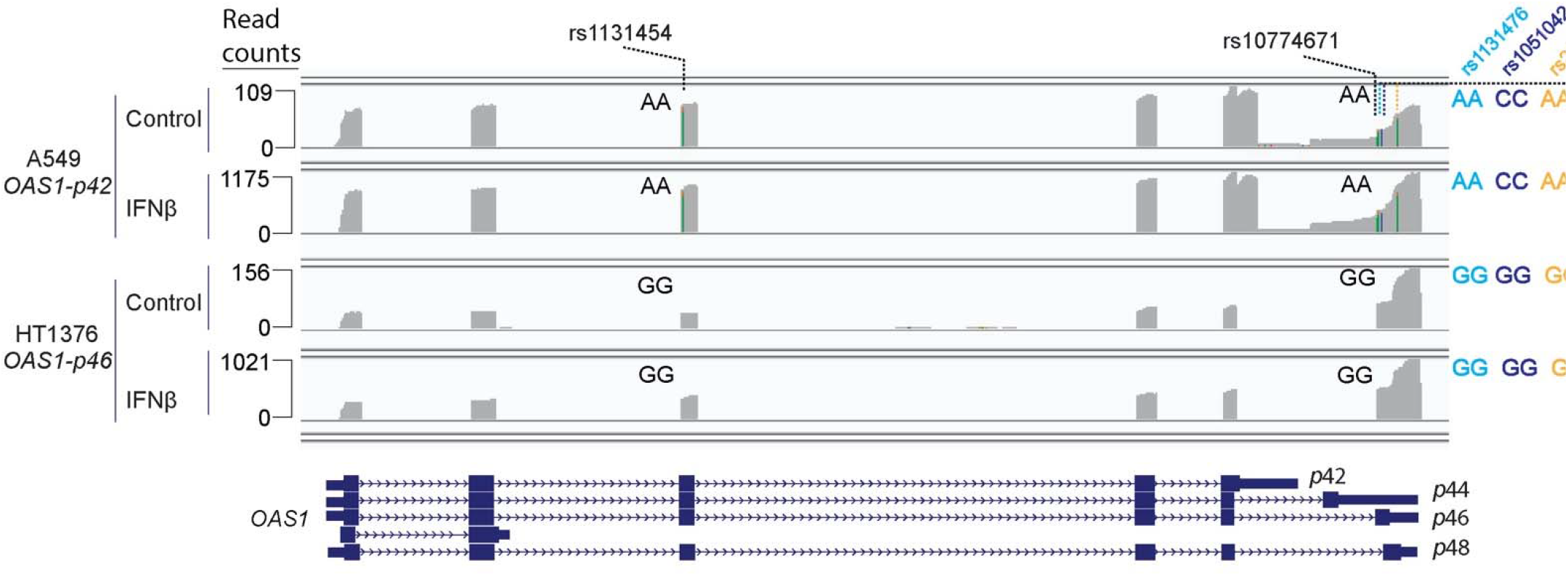
Long-read sequencing of *OAS1* transcripts in A549 and HT1376 cells with and without IFNβ treatment. The A549 (rs10774671-AA, *OAS1-p42*) and HT1376 (rs10774671-GG, *OAS1-p46*) cells were treated with IFNβ or PBS (control) for 48 hrs. Total RNA was harvested and subjected to long-read sequencing with Oxford Nanopore. In both cell lines, the expression of *OAS1* isoforms was strongly induced by IFNβ treatment. The IGV plots show auto-scaled profiles with read counts indicating maximum coverage in the window. Genotypes of transcribed variants are indicated.

**Figure S6.**
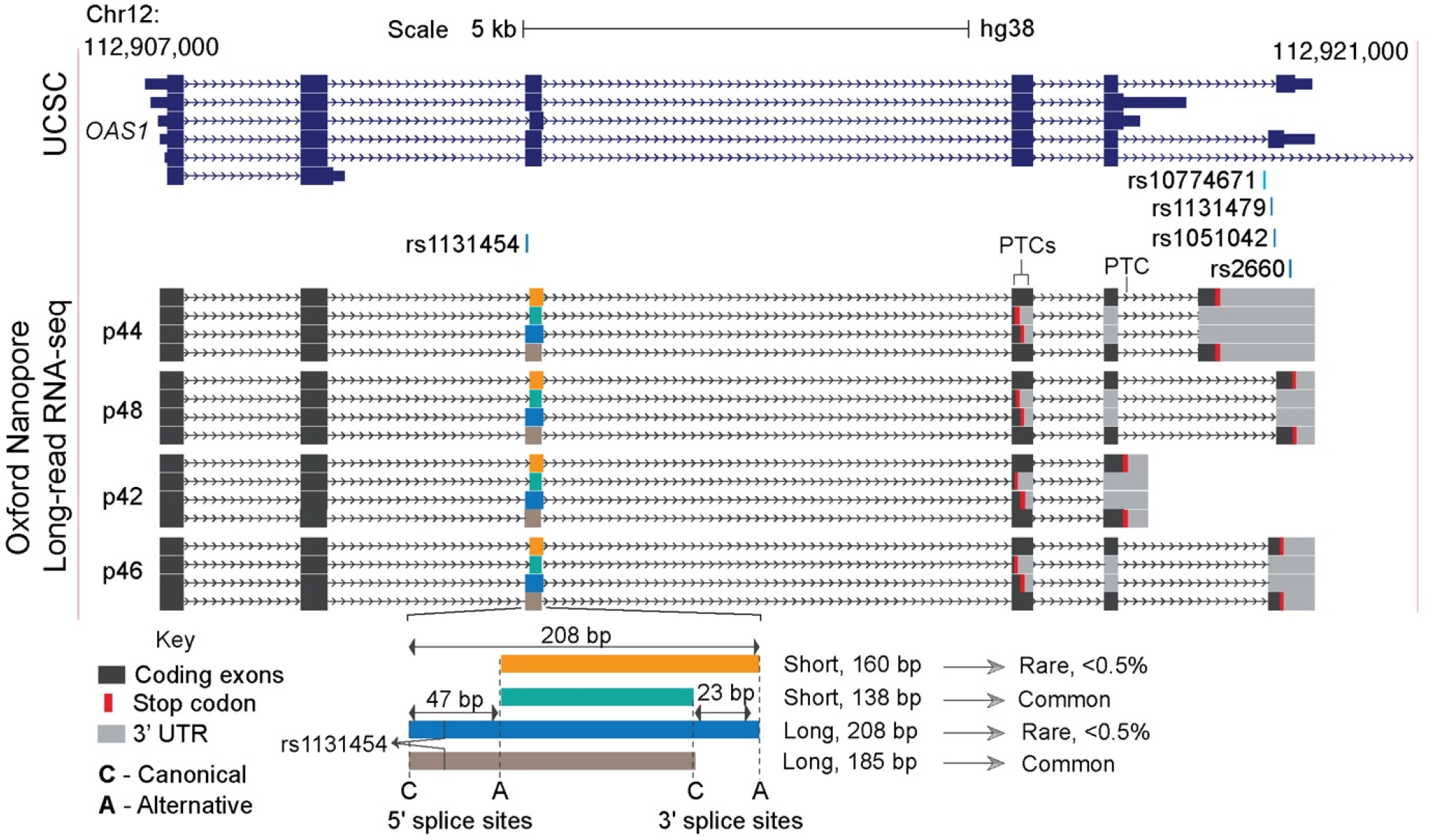
Long-read sequencing of OAS1 transcripts demonstrates functional consequences of alternative splicing of exon 3. UCSC genome browser view shows the known *OAS1* transcripts and full-length sequences of *OAS1* isoforms determined by long-read RNA-seq with Oxford Nanopore (**Figure S5**). Splicing patterns of *OAS1* exon 3 depend on two acceptor and two donor splice sites that define the Short and Long exon 3 isoforms. Two common exon 3 isoforms are Short (138 bp) and Long (185bp), and rare (<5% reads) exon 3 isoforms are Short (160 bp) and Long (208 bp). The inclusion of common Short (138 bp) or rare Long (208 bp) exon 3 isoforms results in premature termination codons (PTC) in exon 4. The stop codon of canonical *p42* isoforms presented by rare Short (160 bp) and common Long (185 bp) isoforms is located within exon 5 and functions as a PTC due to the inclusion of additional exons creating *p44* or *p48* isoforms. All transcripts with PTC are targeted by nonsense-mediated decay (NMD) with a variable extent of degradation depending on specific isoforms.

**Figure S7.**
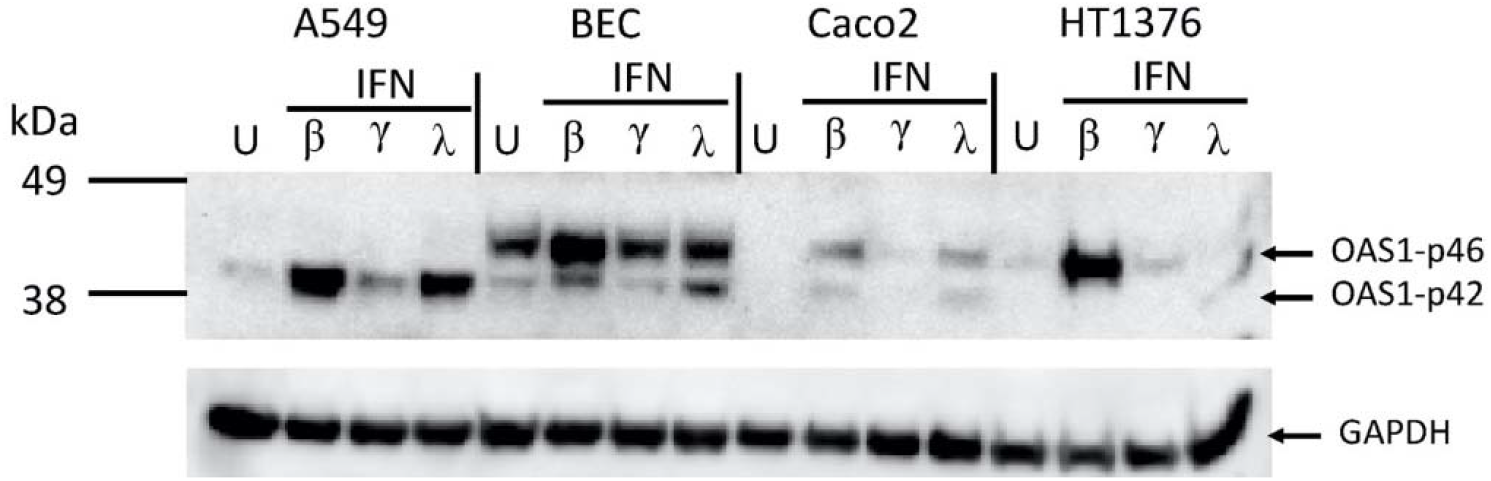
Western blot analysis of expression of endogenous OAS1-p42 and p46 protein isoforms in human cell lines. Four cell lines with different genotypes of rs10774671: A549 (AA), primary bronchial epithelial cells BEC (AG), Caco2 (AG), and HT1376 (GG) were treated with interferons – IFNβ (2ng/mL), IFNγ (2ng/mL) or IFN-λ3 (100 ng/mL) for 48 hrs. Similar amounts of protein lysates were used for Western blot with C-terminal OAS1 antibody. Only two isoforms, OAS1-p42 and p46, were detected, with p46 protein being dominant in heterozygous cell lines. In homozygous cell lines, the protein expression of p42 and p46 appeared comparable after IFN treatment.

**Figure S8.**
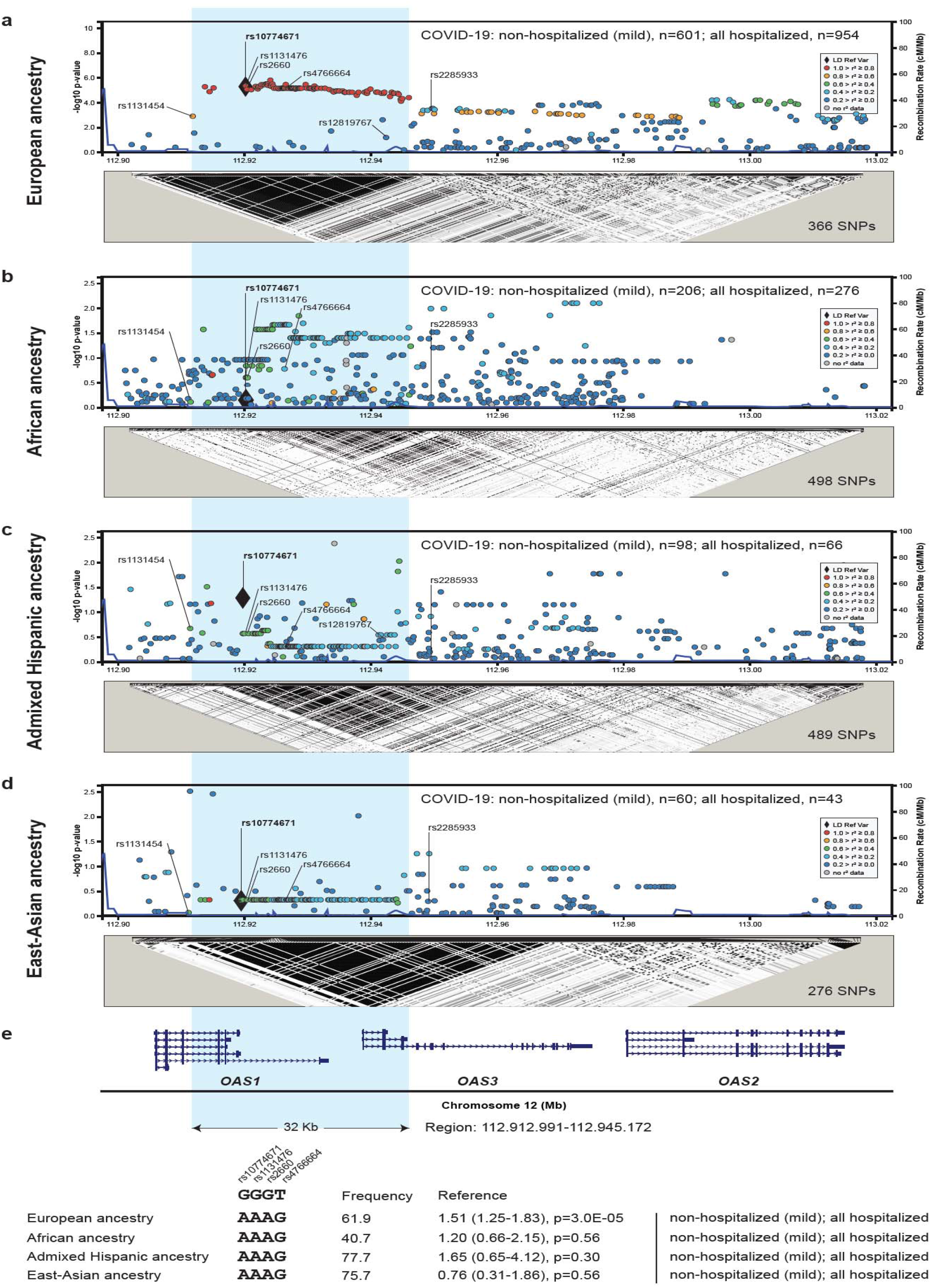
Association analyses within the chr12q24.13 region in relation to COVID-19 outcomes in patients of diverse ancestries. Association analyses and LD plots (r2) in patients with non-hospitalized (mild) vs. hospitalized COVID-19 in patients of **a)** European ancestry (n=1555); **b)** African ancestry (n=483); **c)** Admixed Hispanic ancestry (n=166); and d) East-Asian ancestry (n=103). e) location of *OAS1, OAS3*, and *OAS2* reference transcripts. Haplotype analysis of four core variants within the 32 Kb LD block (hg38: chr12:112912991-112945172), marked by blue shading, tagging the Neandertal haplotype in Europeans. Association results comparing the main risk vs. main non-risk (Neandertal) haplotypes. Full association results for individual variants and haplotypes are provided in **Tables S4-S8**.

